# Comparison of control and transmission of COVID-19 across epidemic waves in Hong Kong: an observational study

**DOI:** 10.1101/2023.06.20.23291593

**Authors:** Bingyi Yang, Yun Lin, Weijia Xiong, Chang Liu, Huizhi Gao, Faith Ho, Jiayi Zhou, Ru Zhang, Jessica Y. Wong, Justin K. Cheung, Eric H. Y. Lau, Tim K. Tsang, Jingyi Xiao, Irene O. L. Wong, Mario Martín-Sánchez, Gabriel M. Leung, Benjamin J. Cowling, Peng Wu

**Author notes:** Corresponding author: Ben Cowling: School of Public Health, Li Ka Shing Faculty of Medicine, The University of Hong Kong, 7 Sassoon Road, Pokfulam, Hong Kong. Funding: Health and Medical Research Fund.

## Abstract

**Background:** Hong Kong contained COVID-19 for two years, but experienced a large epidemic of Omicron BA.2 in early 2022 and endemic transmission of Omicron subvariants thereafter.

**Methods:** We examined the use and impact of pandemic controls in Hong Kong by analysing data on more than 1.7 million confirmed COVID-19 cases and characterizing non-pharmaceutical and pharmaceutical interventions implemented from January 2020 through to 30 December 2022. We estimated the daily effective reproductive number (R_t_) to track changes in transmissibility and effectiveness of community-based measures against infection over time. We examined the temporal changes of pharmaceutical interventions, mortality rate and case-fatality risks (CFRs), particularly among older adults.

**Findings:** Hong Kong experienced four local epidemic waves predominated by the ancestral strain in 2020 and early 2021 and prevented multiple SARS-CoV-2 variants from spreading in the community before 2022. Strict travel-related, case-based, and community-based measures were increasingly tightened in Hong Kong over the first two years of the pandemic. However, even very stringent measures were unable to contain the spread of Omicron BA.2 in Hong Kong. Despite high overall vaccination uptake (>70% with at least two doses), high mortality was observed during the Omicron BA.2 wave due to lower vaccine coverage (42%) among adults ≥65 years of age. Increases in antiviral usage and vaccination uptake over time through 2022 was associated with decreased case fatality risks.

**Interpretation:** Integrated strict measures were able to reduce importation risks and interrupt local transmission to contain COVID-19 transmission and disease burden while awaiting vaccine development and rollout. Increasing coverage of pharmaceutical interventions among high-risk groups reduced infection-related mortality and mitigated the adverse health impact of the pandemic.

## INTRODUCTION

The pandemic of coronavirus disease 2019 (COVID-19) caused by SARS-CoV-2 led to billions of infections, and more than 17 million deaths had been recorded globally from late 2019.^1^ Compared to the most recent influenza pandemic in 2009-2011, significant improvements have been made in pandemic responses including surveillance, diagnostics, and innovation and rapid development of vaccines and antivirals.^2^ Once effective pharmaceuticals became available for SARS-CoV-2, many locations began to transition away from the use of stringent non-pharmaceutical interventions (NPIs) to control transmission, given the substantial social and economic costs of sustaining these measures. Rapid evolution of SARS-CoV-2 resulted in the emergence of new variants, including Omicron that has dominated the largest waves of global transmission.^3,4^ The timing and impact of pandemic waves have varied substantially around the world.^5-7^ Identifying the optimal combination of pharmaceutical and non-pharmaceutical measures at different points in time remains a priority not only for reflecting on the response to the COVID-19 pandemic but also for preparing for the next global outbreak.

As a unique case, Hong Kong adopted a stringent containment strategy since 2020, with successful control of four local epidemic waves in 2020 and the first half of 2021, and no major outbreaks between April and December 2021. However, a large community epidemic of Omicron beginning in January 2022 caused a large number of fatalities within three months, despite the availability of COVID-19 vaccines for over a year by that time, and more stringent NPIs being implemented to attempt to contain transmission.^3,8^ Here, we systematically examine the progression and control of COVID-19 pandemic waves in Hong Kong, focusing on locally adopted control measures and their impact on transmission and burden of COVID-19, to identify critical factors in response and preparedness over the course of the pandemic.

## METHODS

### Sources of data

We obtained demographic, clinical, and epidemiological data (situation as of 29 January 2023) on all laboratory-confirmed and self-reported cases between January 2020 and December 2022 from the Hong Kong Department of Health and Hospital Authority of Hong Kong. Cases, regardless of symptoms, were recorded as laboratory-confirmed when positive by reverse transcription polymerase chain reaction (RT-PCR) tests throughout the pandemic and recorded as confirmed based on self-reported positive rapid antigen tests (RAT) since 26 February 2022. Confirmed COVID-19 cases were classified into mild/moderate, serious, critical, and fatal according to clinical outcomes, while confirmed cases with available epidemiological information were further classified into imported, linked-to-imported, local, and contacts of local cases (Appendix).

We classified the COVID-19 pandemic in Hong Kong into six waves by confirmation date of cases. Each wave was divided into pre-peak (16-50 days before peak), peak (15 days before or after peak), and post-peak periods (16-50 days after peak) to reflect the trajectory, where peak was defined as the day recording the largest number of confirmed cases. Waves 5 and 6 were further divided into two periods to reflect changes in healthcare and predominant virus variant over the epidemic, namely 5a (31 Dec 2021 - 6 Feb 2022), 5b (7 Feb - 22 May 2022), 6a (23 May - 30 Sep 2022) and 6b (1 Oct - 31 Dec 2022). Although the Hong Kong SAR Government classified the resurgence of infections in June 2022 with Omicron BA.4/5 as a continuation of the fifth wave, because daily cases never declined to zero prior to the resurgence, here we refer to this as the sixth wave for clarity and consistency. Our project was approved by the Institutional Review Board of the University of Hong Kong/Hospital Authority Hong Kong West Cluster.

### Characterization of non-pharmaceutical interventions

Information on NPIs implemented in Hong Kong was collected through the government press releases and classified into travel-related, community-wide, and case-based measures (details in Appendix). Travel-related measures include entrance restrictions, inbound traveller testing, quarantine, and exemptions. Community-wide measures include school closures, work-from-home policies, mask wearing, restrictions on group gatherings and measures to reduce crowding and mixing in the population. Case-based interventions refer to targeted measures related to case identification, timely isolation, and tracing and quarantine of contacts.

We used contact tracing data to assess the temporal changes in local case clustering patterns. A cluster was defined as at least two RT-PCR confirmed cases with contact history and initiated by index local cases (Appendix).^9^ We characterized the monthly distribution and density distribution of cluster size, stratified by waves.

### Case finding and contact tracing

We obtained data on COVID-19 RT-PCR tests performed in Hong Kong through various testing schemes (Appendix) up to wave 5a (7 February 2022),^10^ including daily numbers of specimens tested and test positives. The testing schemes focused on different risk populations, including inbound travellers, cases and their close contacts, patients in clinical settings, and persons in the general community. We describe temporal changes in testing capacity and case detection proportions and compare the two-week moving average of numbers of specimens collected from close contacts to that from the confirmed cases. We report the distribution of the delays between illness onset and case confirmation for symptomatic cases as indicators of the potential impact on transmissibility of case finding and isolation across waves, stratified by type of case, wave and trajectory.

### Population behavioural responses to community-based measures

To assess population behavioural responses during the COVID-19 pandemic, we conducted 107 rounds of cross-sectional random digit dialling telephone surveys from January 2020 to December 2022 (details in appendix).^11,12^ Participants reported on face mask usage, personal hygiene, and social distancing behaviours in the week prior to each survey. We calculated the rim-weighted^11^ proportion of respondents who reported specific behaviours in each survey, with binomial confidence intervals (CIs).

We compared community mobility to the pre-pandemic level (1 January 2020) using transaction data on Octopus cards, which is an ubiquitous stored-value and age-stratified payment method for daily public transport.^13^ Overall mobility based on all card types was calculated and weighted by population age-structure. Changes in community mobility were analysed, stratified by time to the peak of each epidemic wave and age group. We also obtained data from the Immigration Department of Hong Kong to assess changes in inbound travellers during the study period.

### COVID-19 transmission and impact of non-pharmaceutical interventions

We extended the approach described by Cori et al.^14^ to estimate the time-varying effective reproductive number (R_t_) for imported and local cases separately to characterise the transmissibility of COVID-19 (Appendix).^15^ We inferred the epidemic curve by infection date for *R_t_* estimation by applying a deconvolution approach^16^ to incubation period (mean 5.2 (SD 3.9) days for waves 1-4 and mean 3.5 (SD 2.6) days for waves 5-6)^17,18^, infectiousness relative to onset time^19^ and infection-to-report delay from empirical data.^12^

We examined the impact of combined NPIs (“NPI package”) on reducing community transmission by monitoring changes in *R_t_* for local cases. The inclusion of individual NPIs was informed by the clustering and the variance explained (Appendix). Log-linear regression models were used with daily *R_t_* as an outcome and interventions as time-varying covariates across waves 1 to 5a, allowing for different initial *R_t_* and effect of work-from-home of each wave. We repeated the above analyses by dropping one wave at each time to test potential changes in NPIs effects across waves. *R_t_* was estimated for defined NPI packages representing different intensities of interventions and the initial condition as reflected by *R_t_*, i.e., the initial transmissibility in each wave, in comparison with the scenarios of no NPIs and applying all available NPIs. 95% CIs were estimated using 1,000 bootstraps of coefficient estimates following a multivariate normal distribution.

### COVID-19 fatality and pharmaceutical interventions

To characterize the changes in disease burden and severity over wave and trajectory, we used the weekly mortality rate and case-fatality risk (CFR) for the overall population, older adults ≥65 years, and unvaccinated individuals, respectively. Within each group and study period, the weekly mortality rate was calculated as the weekly number of COVID-19 deaths over person-times observed, while the CFR was defined as the number of deaths over the total number of confirmed COVID-19 cases (details in appendix). Data from 1 October 2022 onwards were excluded due to potentially reduced ascertainment of COVID-19 cases after that date.

Two vaccines including the inactivated vaccine CoronaVac (Sinovac) and the mRNA vaccine BNT162b2 (BioNTech/Fosun Pharma/Pfizer) were provided for free in Hong Kong starting from February 2021, with third doses available for adults since November 2021. We collected age-specific population and daily numbers of vaccine doses administered by age from the territory-wide vaccine registry to calculate the proportion fully vaccinated (≥2 doses) among the overall population and older adults ≥65 years of age, stratified by wave and trajectory.

Two COVID-specific oral antiviral drugs, molnupiravir (Merck) and nirmatrelvir/ritonavir (Pfizer) have been authorized for use in Hong Kong since late February 2022 and mid-March 2022 respectively. Drug prescriptions for individual patients were used to calculate the proportion of patients who were confirmed by either RT-PCR or RAT and received the antivirals in all ages and unvaccinated older adults ≥65 years, stratified by wave and trajectory.

All analyses were conducted in R version 4.0.0 (R Foundation for Statistical Computing, Vienna, Austria).

## RESULTS

### Non-pharmaceutical interventions and COVID-19 transmission

Hong Kong experienced six epidemic waves from January 2020 through December 2022 (Figure 1, Figure S1 and Table S1), with over 2.6 million laboratory- and RAT-confirmed COVID-19 cases (Table 1). Notwithstanding differential ascertainment by wave, only 12,631 (0.5%) cases were confirmed during the first four epidemic waves predominated by the SARS-CoV-2 ancestral strain, corresponding to 1.6 cases per 1,000 population. In the second half of 2021, in total there were 841 (98%) imported cases but only five sporadic/index local cases reported (Table 1, S2). A superspreading event associated with a case of Omicron BA.2.2 who acquired the infection in a quarantine hotel initiated a large fifth wave, with a cumulative incidence of 162 cases per 1,000 persons during January-May 2022 (wave 5), and the incidence rates dropped by 90% afterwards (Figure 1).

**Figure 1.**
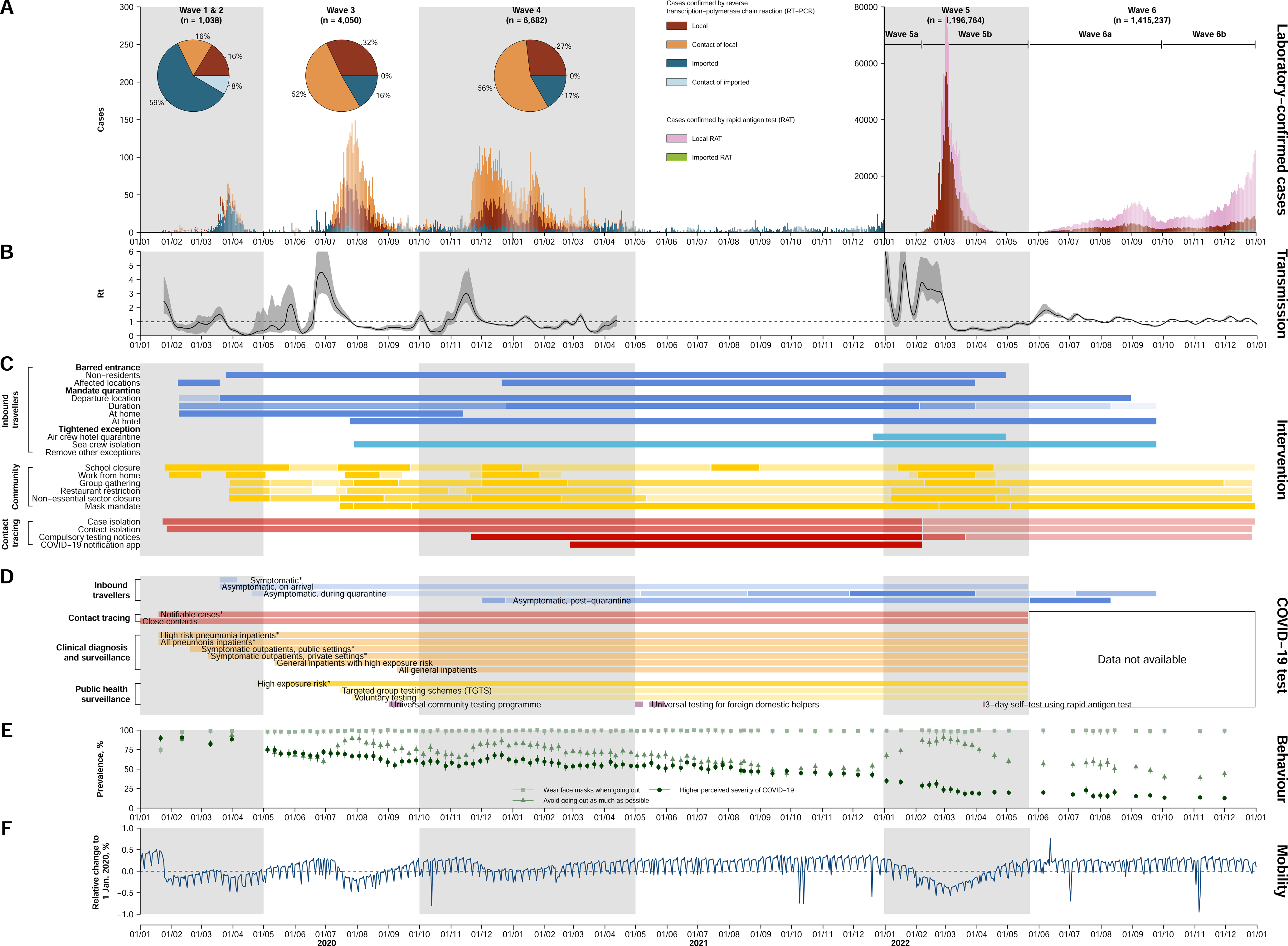
Confirmed cases (A), transmission dynamics (B), non-pharmaceutical interventions (NPIs, C), testing schemes (D), self-reported population behaviours (E), and population mobility (F) across the six epidemic waves of COVID-19 in Hong Kong. (**A**) COVID-19 cases. Coloured bars indicated cases laboratory-confirmed by RT-PCR or self-reported rapid antigen test (RAT) positives, which were only available after February 25, 2022. Laboratory-cases were classified into sporadic/index imported (dark blue), linked-to-imported (light blue), sporadic/index local (brown) and contacts of local COVID-19 cases (orange). Self-reported RAT positive cases were classified into imported (green) or local (pink) cases. Pie charts shows the share of case type for the first four waves. (**B**) Estimated effective reproduction numbers (*R_t_*) for local transmission based on identified local cases. Shaded areas in dark grey indicated the estimated 95% credible intervals of *R_t_*. (**C**) NPIs taken to suppress and contain the Covid-19 transmission in Hong Kong by time. NPIs were classified into three target groups: inbound travellers (blue), community (yellow) and case and contact tracing (red). Darker shading represents more stringent measures, with details in Table S4. (**D**) COVID-19 testing schemes by target groups and settings. Items with asterisks indicate testing for individuals with respiratory illness. (**E**) Population behaviours related to physical distancing and personal hygiene measured among the general adult population across 107 cross-sectional telephone surveys, January 2020 to December 2022. Point estimates (points) and 95% confidence intervals (vertical segments) were estimated by each survey. (**F**) Percentage changes in transport transactions using Octopus cards relative to 1 January 2020, weighted by age-structure of Hong Kong population in 2021.

**Table 1.**
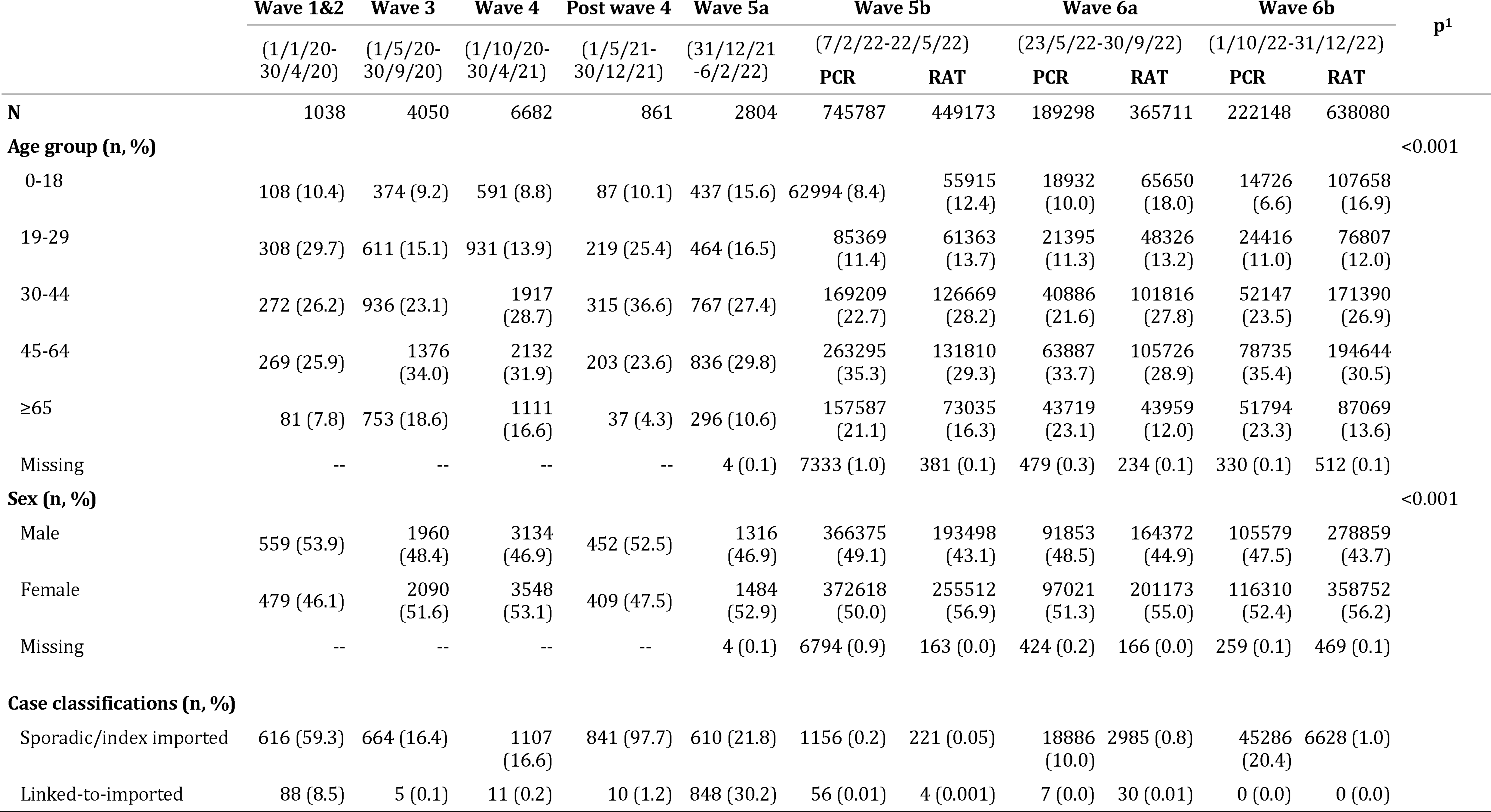
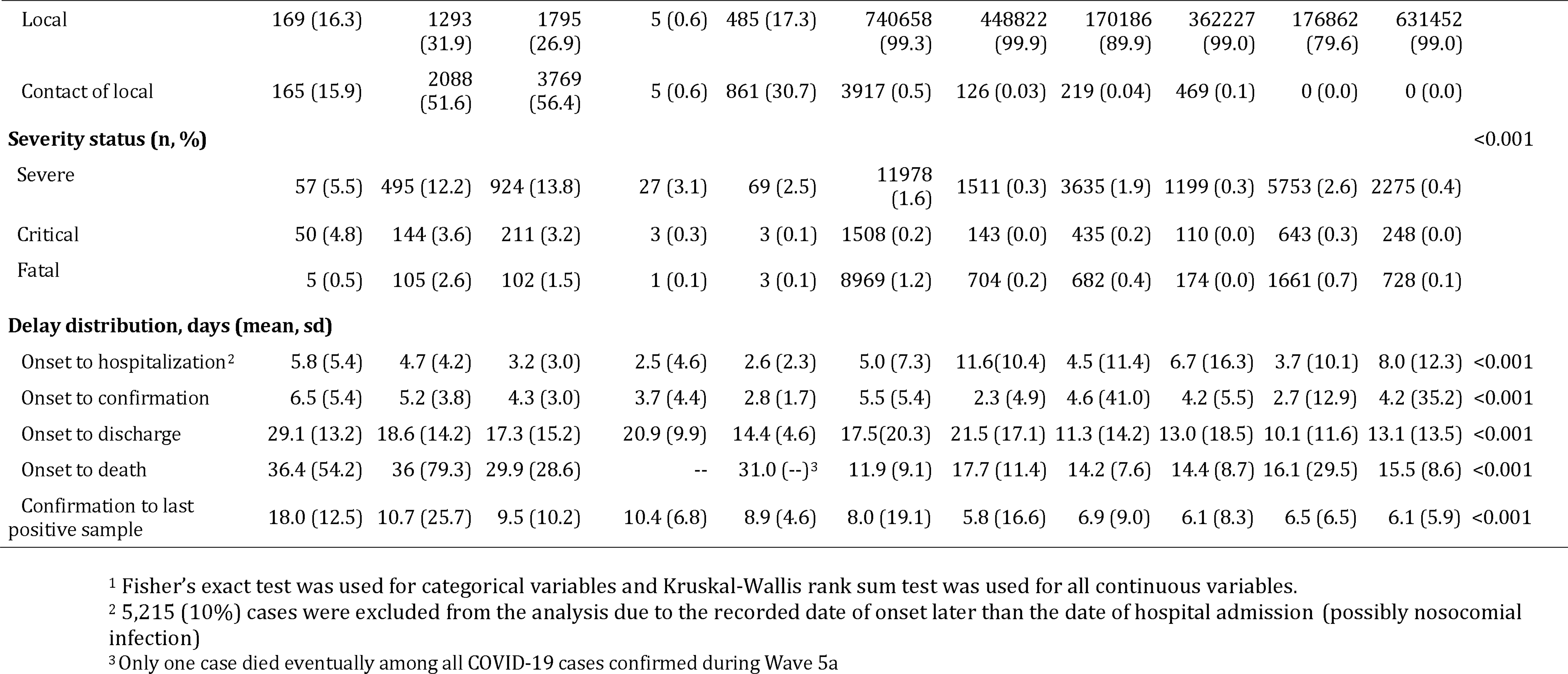
Characteristics of COVID-19 cases confirmed in Hong Kong by epidemic wave.

The estimated *R_t_* for local infections over the epidemic waves (Figure 1B) indicated that transmissibility of SARS-CoV-2 decreased following (re-)implementation of strict NPIs and rose after relaxation of the measures during the first four waves. In the Omicron BA.2.2 wave (wave 5), *R_t_* remained above 1 even after increasingly stringent control measures were implemented during pre-peak and peak periods (wave 5), and mostly fluctuated at around 1 during the Omicron BA.4/5 wave (wave 6).

Hong Kong adopted stringent border controls to prevent importation of cases, including suspending 12 out of 15 boundary control points, barring entry of non-local residents if they have been to overseas places, and mandating on-arrival quarantine with RT-PCR testing in designated facilities for up to 21 days (Figure 1C-D and Table S3-4). We observed a significant decrease in daily passenger arrivals from over 150,000 in January to below 3,000 since April 2020 (a 98% decrease from January 2020) and an increase in daily COVID-19 testing for inbound travellers from 1,000 in March 2020 to 6,000 between July and November 2021 (Figures S2-3). As a result, the *R_t_* for imported cases was maintained well below 1 since mid-February 2020 (Figure S4) and eventually became unquantifiable due to too few sporadic transmission events during the following waves (Table 1).

Hong Kong implemented proactive case finding and contact tracing measures, with substantially expanded laboratory testing capacity over time, leading to gradually shortened onset-to-report intervals (e.g., for close contacts of local cases from a median of 5 to 3 days between wave 3 and 5a) (Figures S5-7, Table S5). Quarantine of close contacts were enforced in designated quarantine facilities since January 2020 and home quarantine was introduced since February 28, 2022, and continued until December 28, 2022. Numbers of specimens collected from close contacts for testing peaked before the fifth wave, but the ratio to confirmed case numbers decreased substantially compared to previous waves (Figure S5), consistent with widespread community transmission. In clinical settings, testing was required for all pneumonia inpatients and outpatients with respiratory illness since February 2020, and expanded to all hospital admissions in May 2020 (Figure 1 and S5). In the community, predefined higher-risk individuals were required to receive regular testing since May 2020, with an increasing contribution to the tested specimens from 40% in the third wave to 88% in wave 5a.

The government closed all schools and recommended civil servants to work remotely in late January 2020 (Figure 1A and Table S2-4). The work-from-home recommendation was removed and re-introduced in subsequent waves, while schools re-opened with restrictions when local cases were low (Figure 1). During the second wave, the government instituted infection control requirements for group gatherings, restaurant capacity and operating hours and other non-essential business sectors in response to local case clusters associated with restaurants and bars. These measures were relaxed and retightened through the following waves. A face mask mandate in all indoor and outdoor public areas was issued since the third wave in mid-July 2020 and remained in place throughout the remaining study period and was only ultimately relaxed in March 2023.

Telephone surveys conducted from late January 2020 and December 2022 (total n=82,562) revealed notable behavioural changes in population response to the implemented NPIs. Physical distancing behaviours were observed at high proportions during waves 1-6 especially when local transmission was high (67% to 90%; Figure S8). Face mask use increased to a high level in January 2020 and remained very high (over 98%) throughout the study period. Reduced behaviour changes and risk perceptions were observed during the period between wave 4 and 5, and during wave 6 (Figures S8-9). There were notable reductions in public transport transactions, with the greatest reductions in wave 5 (largest reduction at 37%, 95% CI, 33% to 40%), compared to previous waves (Figure S10-11). Clusters with more than 10 cases involved were less frequently observed after strict community-based measurements were implemented in the third wave, with a median size of 10 (IQR: 5 to 19) (Figure S12).

We included a combination of NPIs that parsimoniously reflected changes in controlling local transmission in the multivariate analysis (Appendix, Figure S13; adjusted R^2^ 67%), to estimate the effects of different NPI packages. The NPI packages adopted in wave 4 appeared to be the strictest among all the waves (Figure 2A). The initial *R_t_* for waves 1&2, 3, 4 and 5a were estimated to be 1.2 (95% CI: 1.0 to 1.5), 2.6 (95% CI: 1.9 to 3.5), 3.0 (95% CI: 2.3 to 3.9) and 9.3 (95% CI: 6.5 to 13.4), respectively (Figure 3B and Table S6). Applying all locally implemented NPIs at the most intensive level was estimated to bring *R_t_* down to below 1 (e.g., 0.6, 95% CI: 0.4 to 0.9 for wave 3), except for wave 5a (3.1, 95% CI: 1.5 to 5.9), while the analysis on the strictest wave-specific NPI packages showed that only the NPI package used in wave 4 seemed sufficient to control epidemics in waves 1-3, but was only marginally effective during wave 4. However, none of the NPI packages from earlier waves could have sufficiently suppressed Omicron transmission in wave 5 (Figure 2C).

**Figure 2.**
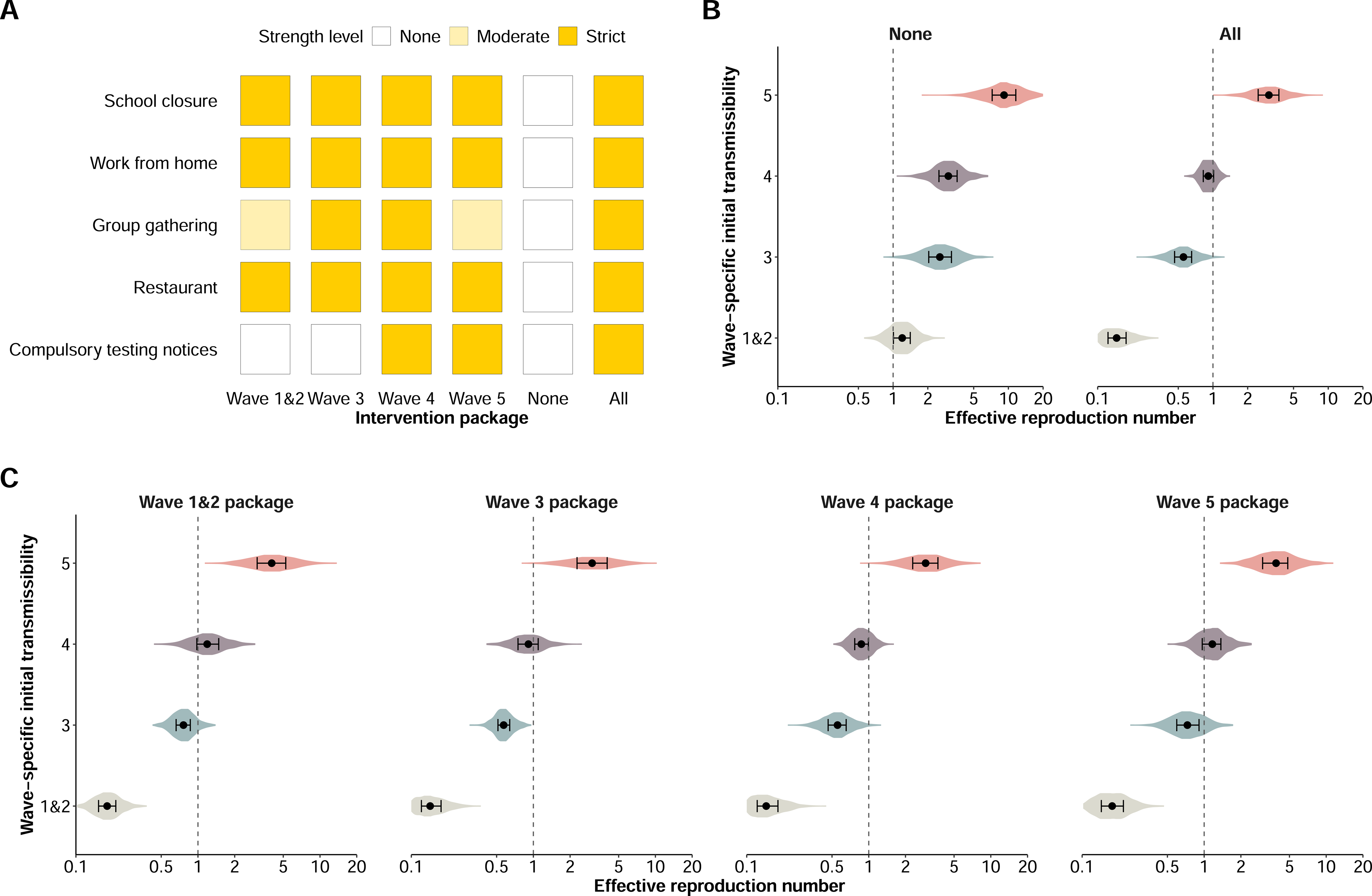
Prediction of effective reproduction numbers (*R_t_*) of COVID-19 associated with non-pharmaceutical interventions (NPIs) taken across epidemic waves. Wave-specific intercepts were used as the baseline level for each epidemic wave. (**A**) NPIs packages used for model prediction: the most stringent measures used in each epidemic wave, no measure, and all measures. (**B**) Wave-specific *R_t_* (row) when no or all measures (column) were implemented. (**C**) Wave-specific *R_t_* (row) under the most stringent NPIs packages (column) of each epidemic wave.

**Figure 3.**
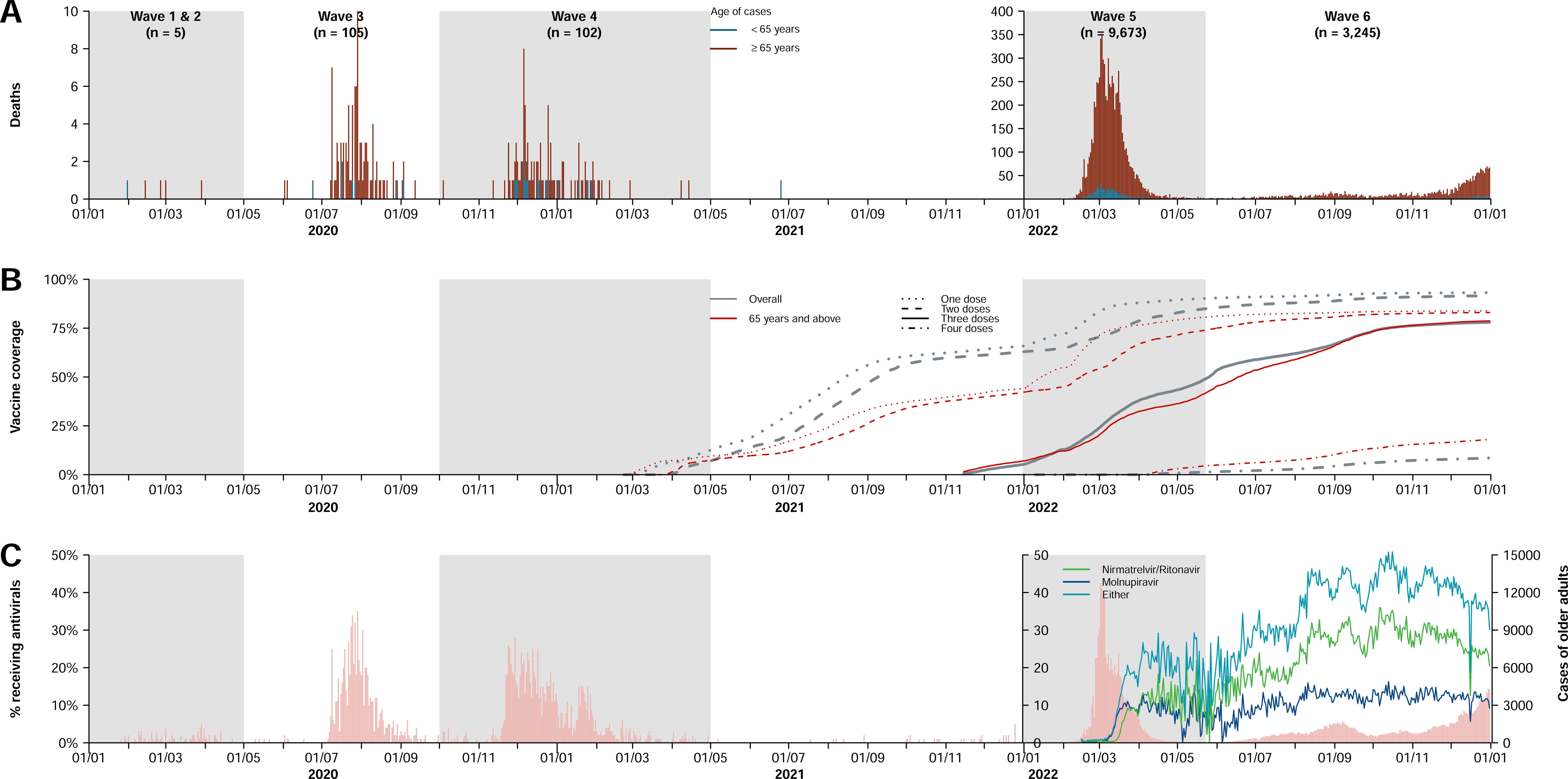
Confirmed deaths, vaccine coverage, and antiviral usages across the six epidemic waves of COVID-19 in Hong Kong. (**A**) Confirmed COVID-19 deaths sorted by date of confirmation. Coloured bars indicated whether the fatal cases were aged 65 years or above (red) or otherwise (blue). (**B**) Cumulative coverage of one, two, three or four doses of CoronaVac and/or BNT162b2. (**C**) Antiviral usage among confirmed COVID-19 cases aged 65 and above sorted by date of confirmation.

### Pharmaceutical interventions against COVID-19

In total 13,134 COVID-19 deaths were reported throughout the six waves, among which 74% (n=9,676) were reported between January and May 2023 (wave 5) with a peak in the overall mortality rate of 2.36 (95% CI, 2.31 to 2.41) deaths per 10,000 person-weeks (Figure 3-4). Among all fatal cases recorded, 92% (n=12,073) were amongst older adults ≥65 years of age.

**Figure 4.**
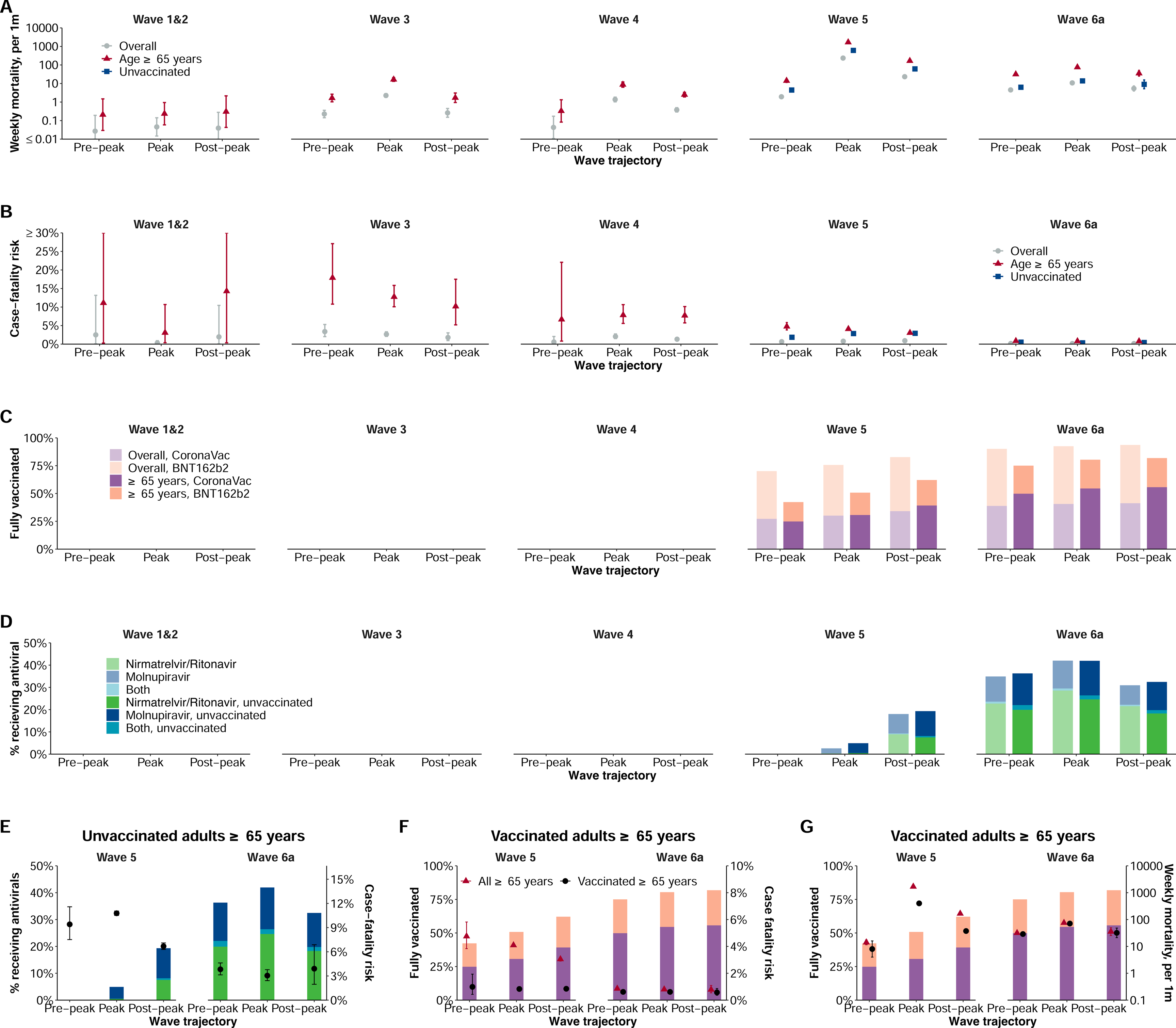
Disease severity and burden of COVID-19 and implementations of pharmaceutical interventions taken across epidemic waves. Estimates were stratified by wave and trajectory. We also calculated the estimates for all Hong Kong population (overall), individuals aged 65y and above, and unvaccinated individuals of all ages (unvaccinated). (**A**) Mortality rate per 1000 persons per week. (**B**) Case-fatality risk. Cases were all notified cases, including PCR and RAT positives. (**C**) Coverage of individuals received two or more doses of COVID-19 vaccines. Vaccine coverage among both all Hong Kong (overall; light colours) and individuals aged ≥65y (dark colours) were calculated. (**D**) Antiviral usage among confirmed COVID-19 cases aged 65 and above. (**E**) Case-fatality risk and antiviral usage in the fifth and sixth wave. Estimates were calculated for unvaccinated individuals aged ≥65y. (**F**) Case-fatality risk and COVID-19 vaccine coverage in the fifth and sixth wave. We calculated vaccine coverage as individuals aged 65 and above and received two or more doses. Case-fatality risk was calculated for all adults ≥65y and fully vaccinated adults ≥65y, respectively. (**G**) Mortality rate and COVID-19 vaccine coverage in the fifth and sixth wave. We calculated vaccine coverage as individuals aged ≥65y who had received two or more doses. Mortality rate was calculated for all adults ≥65y and fully vaccinated adults ≥65y, respectively.

Uptake of the two COVID-19 vaccines in Hong Kong gradually increased during the first six months after having become available, reaching a plateau between October 2021 and January 2022 (Figure 3). Before wave 5, approximately 70% of the population and 42% of older adults ≥65 years received ≥2 doses, but fewer than 10% of the whole population and fewer than one-tenth of older adults had received a third dose (Figures 3-4). The coverage of ≥2 doses of vaccine increased to 82% among adults ≥65 years by the end of wave 6.

The two antivirals molnupiravir and nirmatrelvir/ritonavir first became available only during the peak of wave 5 and were provided to 18% of the adult patients ≥65y within one month (Figures 3-4). The usage of antivirals increased to about 40% among adults ≥65y.

The CFR in infected adults ≥65 years decreased across epidemic waves from 13% (95% CI, 11% to 16%) in wave 3 to 0.83% (95% CI, 0.76% to 0.89%) in wave 6 (Figure 4B). Following availability of the antivirals, CFRs among unvaccinated adults ≥65y decreased from 9.4% (95% CI, 7.5% to 11.6%) pre-peak of wave 5 to 3.0% (95% CI, 2.4% to 3.8%) during the peak of wave 6 (Figure 4E). We observed reduced differences in CFRs and weekly mortality rate between all and vaccinated adults ≥65 years along with the increasing vaccine uptake in this age group (Figure 4F-G).

## DISCUSSION

Using detailed individual case data and territory-wide population data collected during the COVID-19 pandemic in Hong Kong, we were able to systematically examine the progression of epidemics from 2020-2022 in relation to local pandemic responses, including implementation of both NPIs and pharmaceutical interventions over the course of the pandemic.

Stringent NPIs aiming to minimize importation risks and interrupt local transmission chains had been implemented in Hong Kong from early in the pandemic, and successfully controlled multiple epidemic waves in the community caused by the ancestral strain. These measures reduced COVID-19 mortality in a highly susceptible population in the absence of effective pharmaceutical interventions through effectively suppressing the spread of infection.^20^ Nevertheless, the containment strategy relying solely on NPIs may not be sustainable because of the cost and disruption of these measures. Successful suppression of transmission leaves a large population susceptible to infection with newly emerging, immunologically distinct viral strains, and the NPIs would be less effective in preventing pre-symptomatic and superspreading transmissions, and in controlling infections with a relatively higher transmissibility.^19,21^ In some countries, relaxation of NPIs resulted in increased COVID-19 mortality.^1,22^ Achieving high vaccination coverage in the most vulnerable groups, particularly older adults, would minimize COVID-19 mortality after containment measures fail or are relaxed.^23^

Strict travel measures were able to minimize COVID-19 introductions into the community.^24,25^ Despite over 2,000 infections in arriving travellers, only three independent introductions accounted for 90% of the local cases between the second and fourth waves.^26^ The other cornerstone of the approaches to COVID-19 elimination in Hong Kong was the strict isolation of all confirmed cases until viral shedding of the patients reached low levels before discharge, and quarantine of close contacts identified from contact tracing at designated facilities. While isolation and quarantine likely reduced transmission of COVID-19, it is well recognized that many infections in the community were never confirmed, and a number of community epidemics occurred despite intense contact tracing and timely quarantine.^12^ As a consequence, the containment of COVID-19 in Hong Kong cannot be attributed to strict isolation and quarantine alone, while it is clear that moderate social distancing measures were necessary to contain community epidemics with the ancestral strain.

Community-based physical distancing measures were widely adopted in different parts of the world early in the pandemic,^5^ and individual behaviours might have also changed in response to perceived risk.^27^ Our analysis showed that the implementation of packages of physical distancing measures, including school closure, working from home and suspension of large gatherings, etc., correlated with subsequent decline in the effective reproductive number (Figure 2) during epidemics of the ancestral strain but these declines were not sufficient to achieve containment of the more transmissible Omicron subvariants. It was not possible to estimate the impact from individual measures which were often implemented together.

Containment of COVID-19 in Hong Kong allowed vaccination rollout in early 2021 which could provide an opportunity for a transition/exit strategy from the use of NPIs for containment. Despite over 70% of the Hong Kong population being fully vaccinated by the end of 2021, the vaccine uptake among adults ≥65 years was low particularly in adults ≥80 years (approximately 25%) before the Omicron wave. With a high incidence of infections with Omicron, there was a high mortality rate of 1,500 per 1,000,000 persons in 2022, the majority of deaths occurring in unvaccinated older adults. Conversely, locations such as Singapore that only considered containment as a temporary approach prior to reaching high vaccination coverage among vulnerable groups recorded much lower mortality rates in their Omicron waves.^5,7^ If all adults ≥65y were fully vaccinated, a large fraction of deaths in that age group during wave 5 in Hong Kong might have been prevented. The successful control of community transmission for two years in Hong Kong, and the intention to continue with a containment approach regardless of vaccination uptake, ^28^ might have contributed to a lack of urgency in increasing vaccination uptake among vulnerable groups,^11,29^ further exacerbating the low coverage of vaccine in Hong Kong. Risk communication that address the risk perception and urgency of vaccine uptake, along with convenient outreach vaccination programs, could help to improve vaccine uptake in most vulnerable populations.^11^

Planning for “living with the virus” is a challenge for many different locations with different control strategies during the pandemic. Clear objectives in responses are essential for selecting appropriate strategies over the course of the pandemic, and the ultimate goal is always to minimize severe cases and fatalities, and to protect healthcare systems from being overwhelmed. In late 2021, emerging variants such as Delta showing increased immune evasion caused concerns about reduced vaccine-induced protection against infection.^30^ However, evidence suggested that COVID-19 vaccines had remained highly effective in preventing severe and fatal outcomes from infections with the variants,^8,31^ suggesting that vaccination would still likely be an effective way to reduce the severe disease burden even if not preventing infections especially in a population with passive immunity conferred by vaccines.^32-34^ While Omicron was associated with milder disease and lower impact in some locations,^4^ infections may have a similar intrinsic severity to the ancestral virus in persons who have not been vaccinated or previously infected.^35^ Real-time risk assessment of emerging variants remains a challenge.

There are several limitations of our study. First, the investigation of health impact of pandemic control measures did not take into consideration the possible negative effects on economy and quality of life, and the estimates might vary based on local infrastructure and cultural factors. Second, we estimated the initial transmissibility for each epidemic wave, but could not attribute the estimates to viral features or population immunity. However, the increased transmissibility in the fifth wave was likely due to increased transmissibility and immune invasion of Omicron variant, due to the low seroprevalence and vaccination acquired antibodies in previous waves.^32-34^

Experience from Hong Kong has indicated that optimal pandemic control lies in timely and efficient implementation of NPIs, along with a high level of population adherence before pharmaceutical interventions become available. Once vaccines or antivirals can be rolled out, the rationale for continuing to apply disruptive NPIs will gradually weaken. Our findings highlight the value of continuously assessing the level of population immunity against severe disease in the light of viral evolution and the changing availability of pharmaceutical agents. In future pandemics caused by other novel respiratory pathogens, employing NPIs as an initial and temporary strategy can help to contain the spread, providing time to implement more sustainable control measures for high-risk individuals to minimize population mortality and impact on public health.

## Data Availability

All data produced in the present study are available upon reasonable request to the authors

## ACKNOWLEDGMENTS

We thank the Department of Health and the Health Bureau (former Food and Health Bureau) of the Hong Kong SAR Government for providing the data for the analysis, and thank Julie Au and Chloe Chui for technical assistance.

## AUTHOR CONTRIBUTIONS

All authors meet the ICMJE criteria for authorship. The study was conceived by BY, GML, BJC and PW. Data analyses were conducted by BY, YL, CL, HG, FH, JZ, RZ, WX and TKT. BY and PW wrote the first draft of the manuscript, and all authors provided critical review and revision of the text and approved the final version.

