## Appendix for "Comparison of control and transmission of COVID-19 across epidemic waves in Hong Kong: an observational study"

#### Supplementary Tables

Table S1. Dates and periods for trajectory of COVID-19 pandemic waves in Hong Kong.

| Wave | Case peak | Trajectory | | |
|---|---|---|---|---|
| | | Pre-peak | Peak | Post-peak |
| 1 | 9/2/2020 | 1/1/2020 – 24/1/2020 | 25/1/2020 - 24/2/2020 | -- |
| 2 | 27/3/2020 | 1/3/2020 – 11/3/2020 | 12/3/2020 - 11/4/2020 | -- |
| 3 | 30/7/2020 | 10/6/2020 – 14/7/2020 | 15/7/2020 - 14/8/2020 | 15/8/2020 – 18/9/2020 |
| 4 | 29/11/2020 | 10/10/2020 – 13/11/2020 | 14/11/2020 - 14/12/2020 | 15/12/2020 – 18/1/2021 |
| 5 | 3/3/2022 | 12/1/2022 – 15/2/2022 | 16/2/2022 - 18/3/2022 | 19/3/2022 – 22/4/2022 |
| 6a | 8/9/2022 | 20/7/2022 – 23/8/2022 | 24/8/2022- 23/9/2022 | 24/9/2022 – 28/10/2022 |

Table S2. Classification and definition of COVID-19 case.

| Case classification | Definition |
|---|---|
| **Severity status of cases** | |
| Mild/Moderate | Cases who did not fulfil the definition of serious, critical or fatal. |
| Serious | Cases who were either:<br>1) On treatment with dexamethasone (+remdesivir), and/or with either baricitinib or IV tocilizumab during hospitalization;<br>2) Ever being $O_2$ desaturated ($O_2$ desaturation level ≤90%);<br>3) Ever required oxygen supplement of 3 litres per minute or more. Cases who also met the criteria of critical or fatal cases were excluded. |
| Critical | Cases ever admitted to ICU, ever required intubation or extracorporeal membrane oxygenation (ECMO) or in shock. |
| Fatal | A person who died within 28 days of his/her first test positive for SARS-CoV-2, regardless of the underlying cause of death. |
| **Type of cases** | |
| Imported | A case which was considered to acquire infections outside Hong Kong, i.e., visited overseas areas during the incubation period (e.g., 14 or 21 days before onset of symptoms). |
| Linked-to-imported | A case which was first confirmed in a cluster (i.e., imported cluster) of COVID-19 cases directly linked to an index imported case. |
| Local | A case 1) which acquired infection in Hong Kong without travel history and 2) identifiable contact to other confirmed local cases or was first identified in a cluster linked to a local case. |
| Contacts of local | A locally infected case which was identified in a cluster linked to a local case and confirmed later than the index local case of the cluster. |
| **Clustering pattern** | |
| Sporadic | A case without identifiable contact to other confirmed local cases. |
| Index | The case first identified in a cluster. |
| Contact | Cases who were identified in a cluster linked to a local or imported case and confirmed later than the index case of the cluster. |

Table S3. Non-pharmaceutical interventions (NPIs) included in the study.

| Control measure | Description |
|---|---|
| **Inbound traveller** | |
| **Ban entrance** | |
| Non-residents | Non-Hong Kong residents are not permitted to enter Hong Kong if they have been to overseas places. |
| Travellers from countries affected by emerging epidemic or variant | Travellers who have recently stayed in countries/regions affected by emerging variants are restricted from boarding passenger flights for Hong Kong. |
| **Mandate quarantine** | |
| Departure location | Compulsory quarantine is required for inbound travellers from specific or all overseas countries/regions. |
| Quarantine duration* | The minimum duration of compulsory quarantine for all inbound travellers or the strictest compulsory quarantine duration for travellers from countries/regions classified as the highest level of transmission risk. |
| Allow quarantine at home | Compulsory home quarantine for inbound travellers. |
| Mandate quarantine at hotel | Compulsory quarantine in a designated quarantine hotel for inbound travellers. |
| **Tightened existing exemptions** | |
| Compulsory quarantine at vessels for sea crew | Sea crew members for goods vessels must undergo compulsory quarantine in the vessel or a designated hotel if they want to disembark from the vessel. They are only exempted from quarantine if they fulfil certain exemption criteria (e.g., direct point-to-point transport between the ship and airport to sign-off/sign-on the ship). |
| Compulsory hotel self-isolation for air crew | Closed-loop air crew members must undergo self-isolation at a designated hotel until their next duty flight. Non-closed loop air crew may enter local community after self-isolation. |
| **Community-based** | |
| School closure | All kindergartens, primary and secondary schools and private schools in Hong Kong should suspend face-to- |

| | |
|---|---|
| | face classes and all on-campus activities on an all-day or half-day basis. |
| Work-from-home | Special work arrangement for civil servants. Private business was encouraged to follow the work at home arrangements. |
| Group gathering | Group gatherings of more than a specific number of persons in public places are prohibited. |
| Restaurant restrictions | Restrictions on catering premises stipulate the time allowed for dine-in services, the maximum number of persons per table and in the restaurant overall, and later vaccine pass compliance for all persons entering the premises. |
| Non-essential sector closure | Bars and pubs, bathhouse, party rooms, clubs, karaoke establishments, mah-jong-tin kau premises, and cruise ships are required to suspend operation or allowed to operate according to the specified Mode of Operation. |
| Mask mandate | The mandatory mask-wearing requirement stipulates a person must wear a mask all the time when the person is entering or present in a specified indoor or outdoor public place. |
| **Contact-tracing** | |
| Case isolated at hospital | Confirmed cases are required to be isolated at hospital or isolation facility upon testing positive, regardless of their symptoms. |
| Contact quarantined | Close contacts of confirmed cases are required to be quarantined in a quarantine centre or hotel, regardless of their infection status. |
| Compulsory testing notices | The government issues notifications to individuals with pre-defined high risk of exposures, with compulsory nucleic acid tests being required. |
| COVID-19 notification app | Risks of potential exposures and compulsory testing notices notifications were sent through "LeaveHomeSafe" COVID-19 contact tracing and mass surveillance mobile app. |

* Quarantine duration of a particular person is subject to the prevailing quarantine requirements and can be affected by travel history and vaccination. We considered the most stringent duration here.

Table S4. Non-pharmaceutical interventions (NPIs) and their stringency levels used between January 2020 and September 2022 in Hong Kong.

| Control measure | Time period | Stringency level |
|---|---|---|
| **Inbound traveller (except from Mainland and Macao)** | | |
| **Ban entrance** | | |
| Non-residents | 1/1/2020-24/3/2020 | No |
| | 25/3/2020-30/4/2022 | Yes |
| | 1/5/2022-31/12/2022 | No |
| Travellers from countries affected by emerging variant | 1/1/2020-6/2/2020 | No |
| | 7/2/2020-19/3/2020 | Yes |
| | 20/3/2020-20/12/2020 | No |
| | 21/12/2020-31/3/2022 | Yes |
| | 1/4/2022-31/12/2022 | No |
| **Mandate quarantine** | | |
| Departure location | 1/1/2020-7/2/2020 | None |
| | 8/2/2020-18/3/2020 | Affected countries |
| | 19/3/2020-25/9/2022 | All inbound travellers except exempted |
| | 26/9/2022-31/12/2022 | None |
| Quarantine duration* | 1/1/2020-7/2/2020 | None |
| | 8/2/2020-28/12/2020 | 14 days |
| | 29/12/2020-31/3/2022 | 21 days |
| | 1/4/2022-11/8/2022 | 14 or 7 days with two consecutive RAT negatives |
| | 12/8/2022-25/9/2022 | 3 days |
| | 26/9/2022-31/12/2022 | None |
| Allow quarantine at home | 1/1/2020-6/2/2020 | None |
| | 8/2/2020-13/11/2020 | Yes |
| | 14/11/2020-25/9/2022 | No |
| | 26/9/2022-31/12/2022 | None |
| Mandate quarantine at hotel | 1/1/2020-24/7/2020 | No |
| | 25/7/2020-25/9/2022 | Yes |
| | 26/9/2022-31/12/2022 | None |
| **Tightened existing exemptions** | | |
| Compulsory quarantine for sea crew | 1/1/2020-28/7/2020 | No |
| | 29/7/2020-25/9/2022 | Yes |
| | 26/9/2022-31/12/2022 | No |
| Compulsory quarantine at destinated facilities for non- closed loop air crew | 1/1/2020-20/12/2021 | No† |
| | 22/11/2021-30/4/2022 | Yes |
| | 1/5/2022-31/12/2022 | No |
| **Community-based** | | |
| School closure | 1/1/2020-24/1/2020 | Open |
| | 25/1/2020-26/5/2020 | Closed |

| | | |
|---|---|---|
| | 27/5/2020-12/7/2020 | Open half-day |
| | 13/7/2020-22/9/2020 | Closed |
| | 23/9/2020-1/12/2020 | Open half-day |
| | 2/12/2020-10/1/2021 | Closed |
| | 11/1/2021-14/7/2021 | Open half-day |
| | 15/7/2021-31/8/2021 | Closed |
| | 1/9/2021-13/1/2022 | Open half-day |
| | 14/1/2022-18/4/2022 | Closed |
| | 19/4/2022-31/12/2022 | Open with daily RAT |
| Work-from-home | 1/1/2020-28/1/2020 | No |
| | 29/1/2020-1/3/2020 | Except for essential service |
| | 2/3/2020-24/3/2020 | No |
| | 25/3/2020-3/5/2020 | Except for essential service |
| | 4/5/2020-19/7/2020 | No |
| | 20/7/2020-23/8/2020 | Except for essential service |
| | 24/8/2020-19/11/2020 | No |
| | 20/11/2020-1/12/2020 | Flexible work |
| | 2/12/2020--17/1/2021 | Except for essential service |
| | 18/1/2021-24/1/2022 | No |
| | 25/1/2022-3/2/2022 | Flexible work |
| | 4/2/2022-31/3/2022 | Except for essential service |
| | 1/4/2022-20/4/2022 | Flexible work |
| | 21/4/2022-31/12/2022 | No |
| Group gathering | 1/1/2020-28/3/2020 | No |
| | 29/3/2020-7/5/2020 | Maximum 4 |
| | 8/5/2020-18/6/2020 | Maximum 8 |
| | 19/6/2020-14/7/2020 | Maximum 50 |
| | 15/7/2020-28/7/2020 | Maximum 4 |
| | 29/7/2020-10/9/2020 | Maximum 2 |
| | 11/9/2020-1/12/2020 | Maximum 4 |
| | 2/12/2020-23/2/2021 | Maximum 2 |
| | 24/2/2021-9/2/2022 | Maximum 4 |
| | 10/2/2022-20/4/2022 | Maximum 2 |
| | 21/4/2022-30/11/2022 | Maximum 4 |
| | 1/12/2022-28/12/2022 | Maximum 12 |
| Restaurant restriction | 1/1/2020-27/3/2020 | No |
| | 28/3/2020-7/5/2020 | ≤ 4 persons per table |
| | 8/5/2020-18/6/2020 | > 4 persons per table |
| | 19/6/2020-10/7/2020 | No |
| | 11/7/2020-21/7/2020 | > 4 persons per table |
| | 22/7/2020-29/10/2020 | ≤ 4 persons per table |
| | 30/10/2020-15/11/2020 | > 4 persons per table |
| | 16/11/2020-28/4/2021 | ≤ 4 persons per table |

| | | |
|---|---|---|
| | 29/4/2021-6/1/2022 | > 4 persons per table |
| | 7/1/2022-4/5/2022 | ≤ 4 persons per table |
| | 5/5/2022-28/12/2022 | > 4 persons per table |
| Non-essential sector closure | 1/1/2020-27/3/2020 | No |
| | 28/3/2020-7/5/2020 | Closed |
| | 8/5/2020-14/7/2020 | Partially open |
| | 15/7/2020-27/8/2020 | Closed |
| | 28/8/2020-21/11/2020 | Partially open |
| | 22/11/2020-17/2/2021 | Closed |
| | 18/2/2021-12/5/2021 | Partially open |
| | 13/5/2021-6/1/2022 | Mostly open |
| | 7/1/2022-20/4/2022 | Closed |
| | 21/4/2022-28/12/2022 | Mostly open |
| Mask mandate | 1/1/2020-14/7/2020 | No |
| | 15/7/2020-31/12/2022 | Yes |
| **Contact-tracing** | | |
| Case isolation | 1/1/2020-22/1/2020 | No |
| | 23/1/2020-7/2/2022 | Mandate hospital isolation |
| | 8/2/2022-31/12/2022 | Hospital/isolation facility/home |
| Contact quarantine | | |
| | 1/1/2020-7/2/2022 | Camp |
| | 8/2/2022-28/12/2022 | Camp/home |
| Compulsory testing notices | 1/1/2020-20/11/2020 | No |
| | 21/11/2020-7/2/2022 | All places with known cases |
| | 8/2/2022-21/3/2022 | Partial places with known cases |
| | 22/3/2022-28/12/2022 | Partial places with known outbreaks |
| COVID-19 notification app | 1/1/2020-25/2/2021 | No |
| | 26/2/2021-7/2/2022 | Yes |
| | 8/2/2022-28/12/2022 | No |

* Quarantine duration of a particular person is subject to the prevailing quarantine requirements and can be affected by travel history and vaccination. We considered the least stringent duration here.

† Air crew who have visited UK/South Africa are subject to self-isolation at hotel for 21 days since 25/12/2020.

Table S5. Laboratory testing schemes

| Source of specimens | Target |
|---|---|
| Probable cases | Patients with respiratory symptoms who had travel history to a place with active community transmission of COVID-19 or close contact with a confirmed case within 14 days before onset of symptoms. |
| Close contacts | Individuals who were in quarantine camps or contact tracing. |
| Targeted group testing scheme | Individuals who were in presence of specified premises with recently confirmed cases or individuals with pre-defined high risks of infections or exposure. |
| Inpatients – pneumonia | Inpatients presenting with pneumonia. |
| Inpatients – general | Screen testing before hospital admission, regardless of the presence of respiratory symptom status. |
| Outpatients | Outpatients with respiratory symptoms. |
| Inbound travellers | Individuals who travelled to Hong Kong from outside, regardless of the presence or absence of respiratory symptoms. |

Table S6. Impact estimates of non-pharmaceutical interventions (NPIs) on COVID 19 transmission and temporal changes across pandemic waves.

| | Complete model | Wave excluded in the model | | | |
|---|---|---|---|---|---|
| | | **Wave 1&2** | **Wave 3** | **Wave 4** | **Wave 5a** |
| **Wave-specific intercept** | | | | | |
| Wave 1&2 | 1.2 (1.0, 1.5) | -- | 1.1 (0.9, 1.5) | 1.1 (0.9, 1.4) | 1.4 * (1.1, 1.7) |
| Wave 3 | 2.6 *** (1.9, 3.5) | 3.4 *** (2.8, 4.1) | -- | 2.5 *** (1.8, 3.4) | 2.8 *** (2.0, 3.8) |
| Wave 4 | 3.0 *** (2.3, 3.9) | 4.1 *** (3.0, 5.6) | 2.9 *** (2.2, 3.8) | -- | 3.0 *** (2.3, 3.9) |
| Wave 5 | 9.3 *** (6.5, 13.4) | 12.3 *** (8.5, 17.8) | 8.5 *** (5.7, 12.6) | 8.9 *** (6.5, 12.0) | -- |
| **School closure** | | | | | |
| Open | Ref. | Ref. | Ref. | Ref. | Ref. |
| Closed | -4.4 (-21.6, 10.4) | -8.0 (-25.6, 7.1) | -11.8 (-39.2, 10.2) | -13.4 (-36.3, 5.6) | 8.7 (-8.2, 23.0) |
| **Group gathering** | | | | | |
| No restriction | Ref. | Ref. | Ref. | Ref. | Ref. |
| Maximum 50 | -27.5 (-82.3, 10.8) | -- | -- | -32.5 (-89.7, 7.4) | -17.7 (-69.4, 18.3) |
| Maximum 8 | 63.0 *** (36.5, 78.5) | 64.1 *** (42.9, 77.4) | -- | 60.9 *** (33.4, 77.1) | 66.0 *** (41.1, 80.4) |
| Maximum 4 | 25.7 (-23.2, 55.2) | 26.0 (-22.2, 55.2) | 68.3 * (17.6, 87.8) | 29.3 (-18.7, 57.9) | 26.6 (-23.0, 56.2) |
| Maximum 2 | 44.5 * (5.1, 67.6) | 44.7 * (5.8, 67.5) | 77.9 ** (41.1, 91.7) | 41.2 (-4.9, 67.1) | 46.4 * (6.9, 69.2) |
| **Compulsory testing notice** | | | | | |
| No | Ref. | Ref. | Ref. | Ref. | Ref. |
| Yes | 4.1 (-15.7, 20.5) | 2.8 (-16.6, 19.0) | 1.3 (-20.4, 19.1) | -- | 2.2 (-18.6, 19.4) |
| **Restaurant** | | | | | |
| No restriction | Ref. | Ref. | Ref. | Ref. | Ref. |
| Restricted dine-in II | 27.5 (-13.2, 53.5) | 44.5 * (9.7, 65.9) | -84.5 (-392.7, 30.9) | 31.4 (-6.4, 55.8) | 22.7 (-21.6, 50.8) |
| Restricted dine-in I | 66.4 *** (46.3, 79.0) | 75.3 *** (58.3, 85.4) | 19.4 (-108.8, 68.9) | 64.6 *** (42.5, 78.2) | 66.0 *** (45.1, 78.9) |
| **Work-from-home** | | | | | |
| No recommendation | Ref. | Ref. | Ref. | Ref. | Ref. |
| Flexible work | | | | | |
| *Wave 3* | -5.9 (-44.3, 22.2) | -5.5 (-42.3, 21.8) | -- | 7.6 (-28.8, 33.7) | -13.8 (-56.5, 17.3) |
| *Wave 4* | -39.7 ** (-73.9, -12.3) | -39.7 ** (-72.7, -13.0) | -45.1 ** (-84.2, -14.4) | -- | -41.5 ** (-77.0, -13.1) |
| *Wave 5a* | 16.3 (-19.7, 41.5) | 17.0 (-17.4, 41.3) | 18.2 (-19.1, 43.7) | 22.1 (-11.4, 45.5) | -- |
| Essential work | | | | | |
| *Wave 1&2* | 39.3 *** (23.7, 51.8) | -- | 40.9 *** (25.1, 53.4) | 39.6 *** (24.3, 51.8) | 39.1 *** (23.1, 51.7) |
| *Wave 3* | -15.9 (-51.8, 11.5) | -15.2 (-49.6, 11.3) | -- | -3.1 (-37.6, 22.7) | -23.8 (-63.5, 6.3) |
| *Wave 4* | -60.0 *** (-106.5, -23.9) | -55.9 *** (-99.8, -21.7) | -62.3 ** (-122.0, -18.7) | -- | -79.3 *** (-134.6, -37.1) |
| *Wave 5a* | -73.1 ** (-145.2, -22.2) | -71.5 ** (-140.3, -22.4) | -75.4 ** (-154.9, -20.7) | -52.6 * (-117.9, -6.9) | -- |
| **Observations** | 492 | 394 | 339 | 297 | 446 |
| **Adjusted $R^2$** | 0.673 | 0.611 | 0.675 | 0.765 | 0.632 |

** p<0.05*

Table S7. Estimated COVID-19 vaccine-preventable deaths in adults aged 65 and over.

| | Wave 5 | | | Wave 6a | | |
|---|---|---|---|---|---|---|
| | Pre-peak | Peak | Post-peak | Pre-peak | Peak | Post-peak |
| **Observation** | | | | | | |
| **Death number** | | | | | | |
| All adults ≥ 65y | 89 | 7139 | 1666 | 364 | 321 | 34 |
| Unvaccinated adults ≥ 65y | 79 | 5378 | 1002 | 104 | 79 | 11 |
| Vaccinated adults ≥ 65y | 8 | 775 | 236 | 238 | 224 | 23 |
| **Case-fatality risk, %** | | | | | | |
| All adults ≥65y | 4.8 (3.8, 5.8) | 4.1 (4.0, 4.2) | 3.1 (2.9, 3.2) | 0.9 (0.8, 0.9) | 0.8 (0.7, 0.9) | 0.8 (0.5, 1.1) |
| Unvaccinated adults ≥65y | 9.4 (7.5, 11.6) | 10.8 (10.5, 11.1) | 6.7 (6.3, 7.1) | 3.8 (3.1, 4.6) | 3.0 (2.4, 3.8) | 3.9 (2.0, 6.9) |
| Vaccinated adults ≥65y | 1.0 (0.4, 1.9) | 0.8 (0.8, 0.9) | 0.8 (0.7, 1.0) | 0.6 (0.5, 0.7) | 0.6 (0.5, 0.7) | 0.6 (0.4, 0.9) |
| **Counterfactual†** | | | | | | |
| **Averted deaths** | 71 (60, 76) | 5689 (5640, 5736) | 1204 (1174, 1232) | 103 (100, 106) | 76 (73, 78) | 9 (8, 10) |
| **Reduction in deaths, %** | | | | | | |
| All adults ≥65y | 79.8 (85.4, 67.4) | 79.7 (80.3, 79.0) | 72.3 (73.9, 70.5) | 28.3 (29.1, 27.5) | 23.7 (24.3, 22.7) | 26.5 (29.4, 23.5) |
| Unvaccinated adults ≥65y | 87.7 (74.1, 93.8) | 89.4 (88.6, 90.1) | 84.2 (82.1, 86.2) | 81.7 (79.4, 84.1) | 78.4 (75.3, 80.4) | 81.8 (72.7, 90.9) |

†In the counterfactual analysis, all cases who were ≥ 65y and not fully vaccinated (i.e., < 2 doses) were assumed with the CFRs that were observed among vaccinated adults ≥ 65y during each wave and trajectory. The number of averted deaths were calculated as the difference between the observed and estimated deaths among who were ≥ 65y and not fully vaccinated.

#### Supplementary Figures

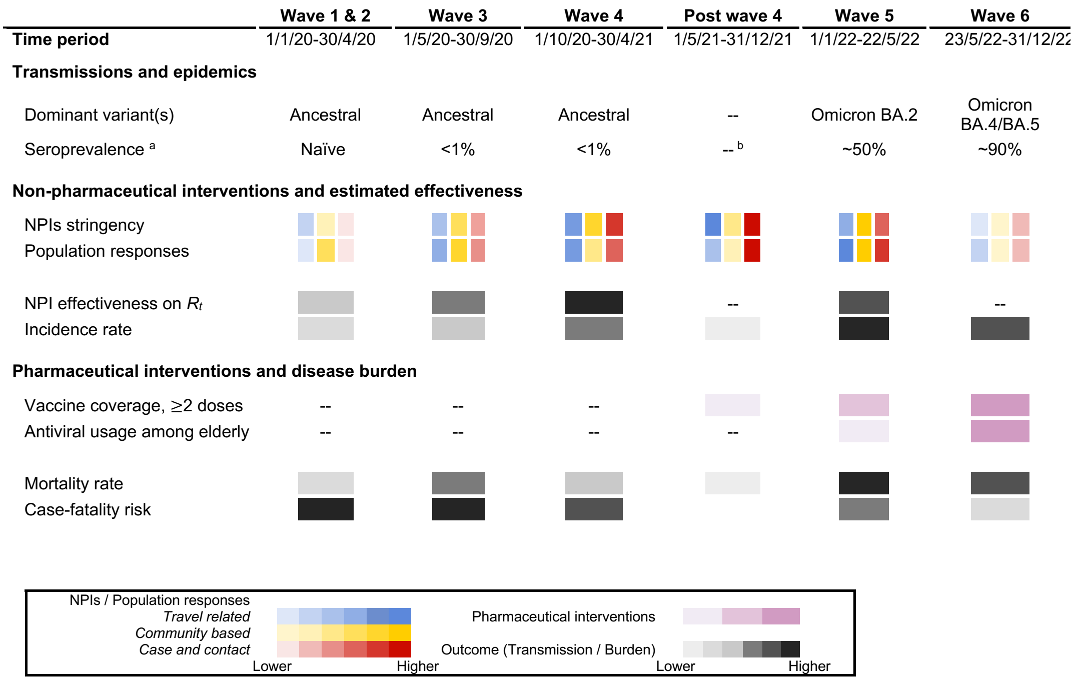

**Figure S1.** Summary of non-pharmaceutical and pharmaceutical interventions, population responses and transmission and diseases burden of COVID-19 across waves in Hong Kong. Darker colour indicates stricter implementations of NPIs; higher level of population responses, disease severity and disease burden; and greater effectiveness in reducing $R_t$, determined by relative ranking among the six epidemic periods (details in Appendix). [a] Seroprevalence estimates at beginning of each wave are obtained from To 2020,[1] Boon 2021[2] and Poon 2022,[3] reflecting antibodies from infections and vaccinations. [b] Seroprevalence at beginning of May 2021 is not available.

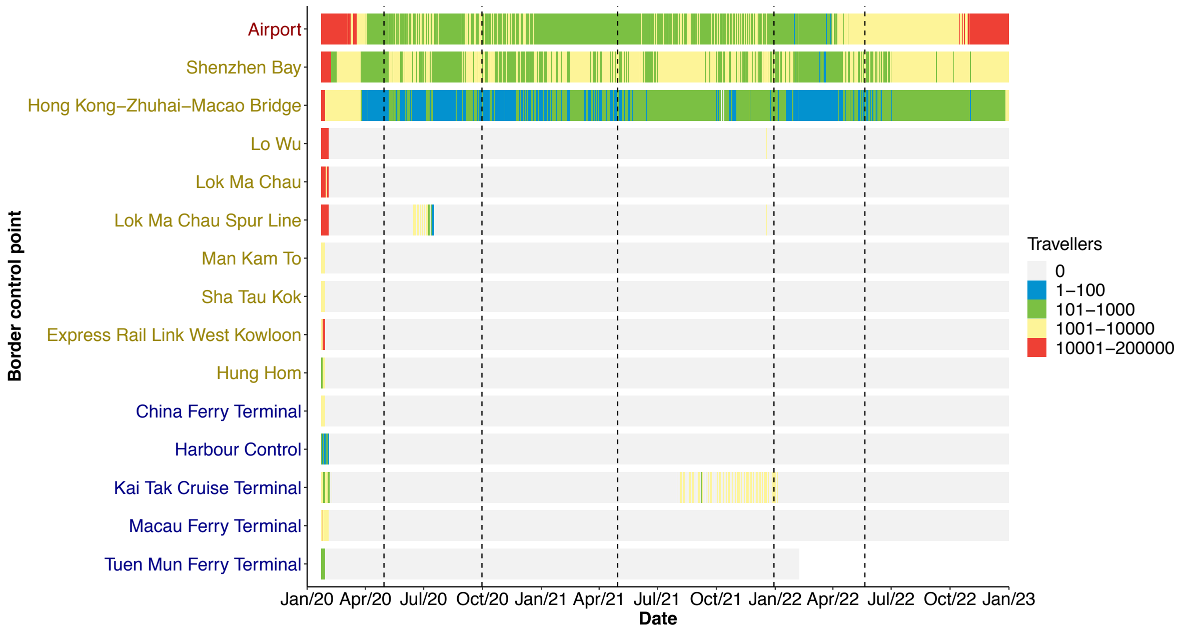

**Figure S2.** Daily number of passengers arriving at Hong Kong through different control points. Red, yellow, and blue indicates control points covering passengers travelling via air, land, and sea. Passengers arriving via land- and terminal-based control points mostly travelled from mainland China and Macao. The COVID-19 pandemic was divided into wave 1 and 2, wave 3, wave 4, post wave 4, wave 5 and wave 6, by vertical dashed lines.

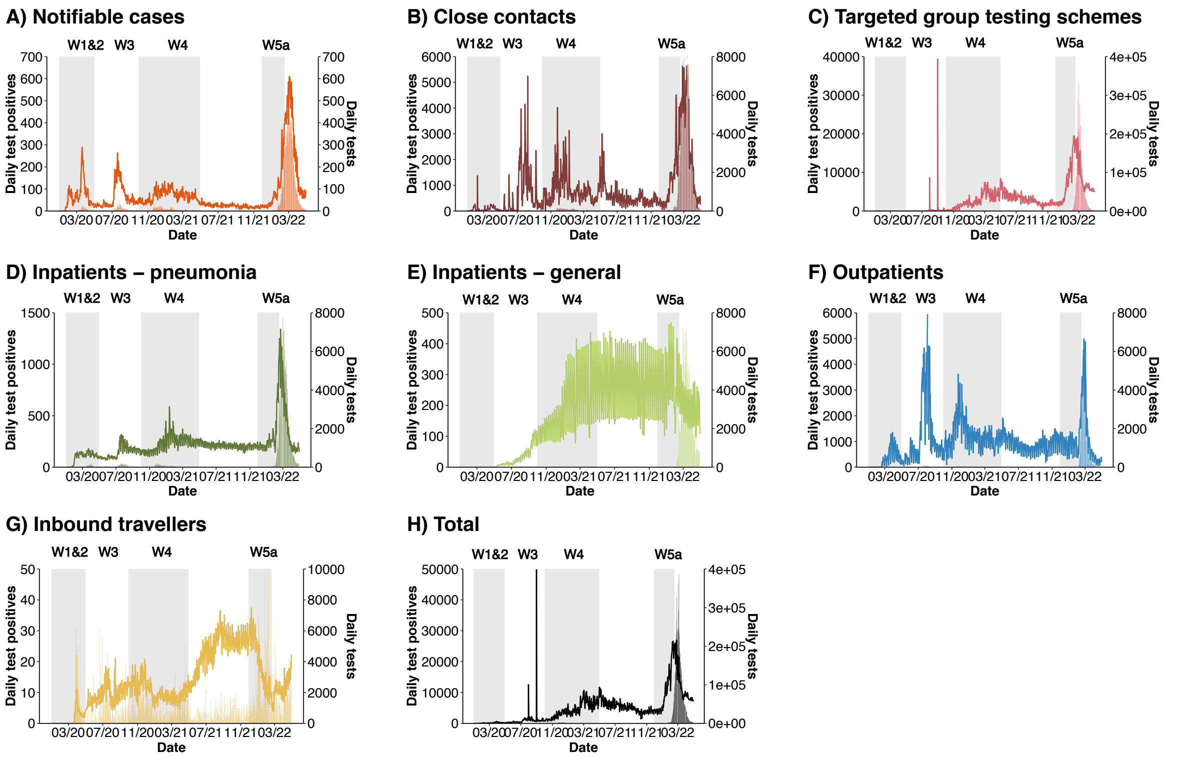

**Figure S3.** Daily COVID-19 test rate (lines) and daily number of test positives (bars) by source of specimen collections. The COVID-19 pandemic was divided into wave 1 and 2, wave 3, wave 4, post wave 4 and wave 5, by grey shades.

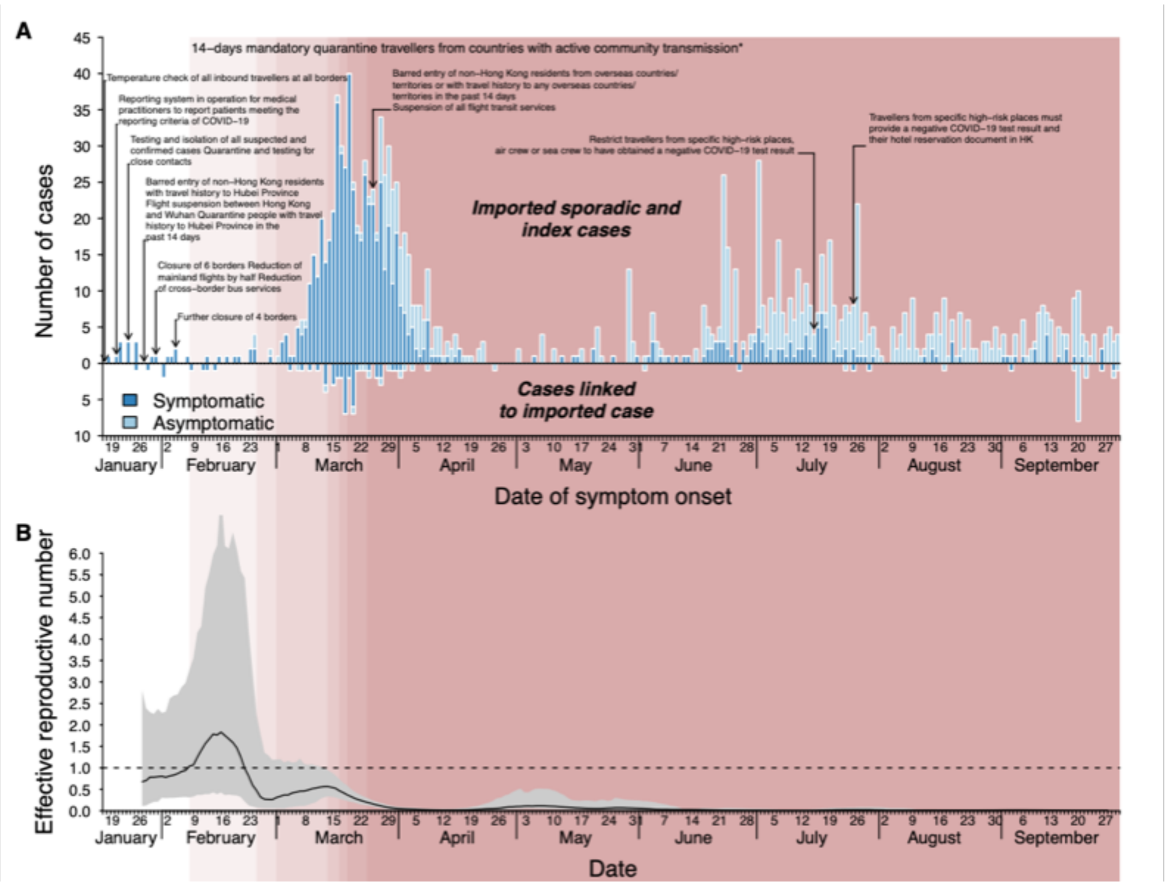

**Figure S4.** Imported COVID-19 cases and estimated effective reproduction numbers ($R_t$) for infections of imported cases across the first three waves in Hong Kong. (A) Sporadic imported COVID-19 cases and index cases linked to imported COVID-19 cases (above), and other cases linked to imported cases (below), by date of symptom onset for symptomatic infections and by date of confirmation for asymptomatic infections. (B) Estimated effective reproduction numbers ($R_t$) for infections of imported cases in Hong Kong. The shaded areas in red indicate the implementation of 14-day mandatory quarantine for all inbound travellers from locations with active community transmission of COVID-19, with darker colours showing the expansion of quarantine order targeting travellers from wider geographic areas.

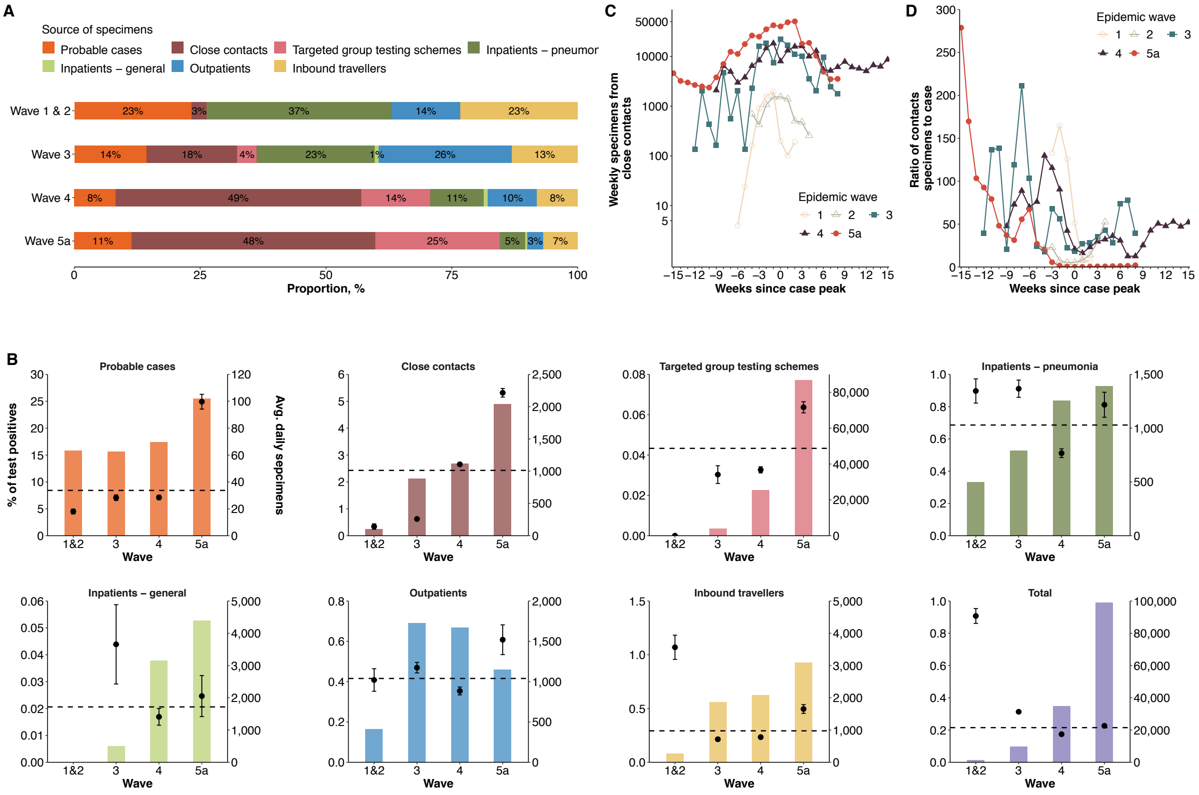

**Figure S5.** Comparison of COVID-19 testing schemes across different waves in Hong Kong. (**A**) Distribution of source of specimen collections among test positives by epidemic wave. (**B**) Average daily number of specimens (bars) tested and percentage of test positives (points and segments), stratified by source of specimen collection and epidemic wave. Horizontal dashed lines indicate the average percentage of test positives by source of specimen collection. (**C**) Weekly number of specimens that were collected from close contacts of confirmed COVID-19 cases relative to case peak of each epidemic wave. (**D**) Moving average of weekly ratio of number of specimens collected from close contacts to the number of confirmed COVID-19 cases relative to case peak of each wave.

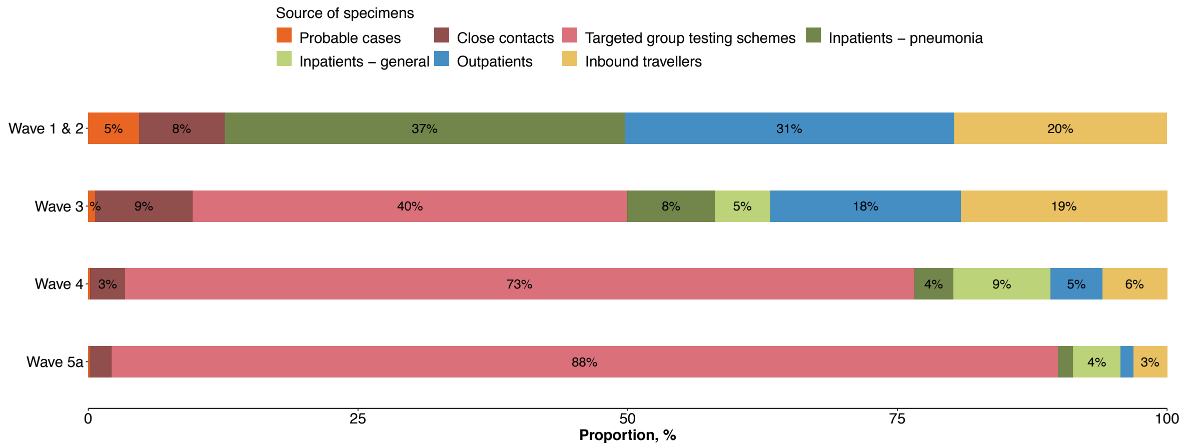

**Figure S6.** Distribution of source of collections among all tested specimens for COVID-19 across different waves.

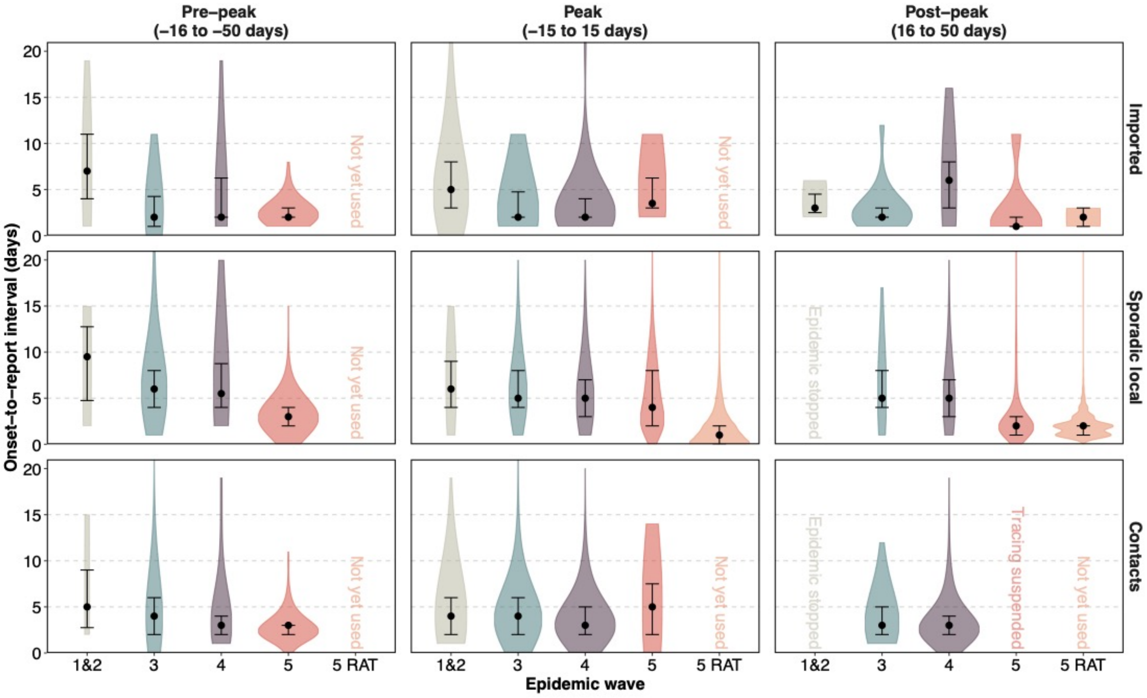

**Figure S7.** Time delays from symptom onset to identification of sporadic imported cases (upper), sporadic/index local cases (middle) and contacts of local cases (lower) across five epidemic waves in Hong Kong. Each wave was divided into three periods, i.e., pre-peak (16-50 days prior to case number peak), peak (case number peak ± 15 days) and post-peak (16-50 days after case number peak). Cases were laboratory-confirmed; otherwise indicated with RAT. No data of local cases and contacts were available for post-peak of wave 1 and 2 due to short epidemic duration. Dots and bars indicate median and interquartile.

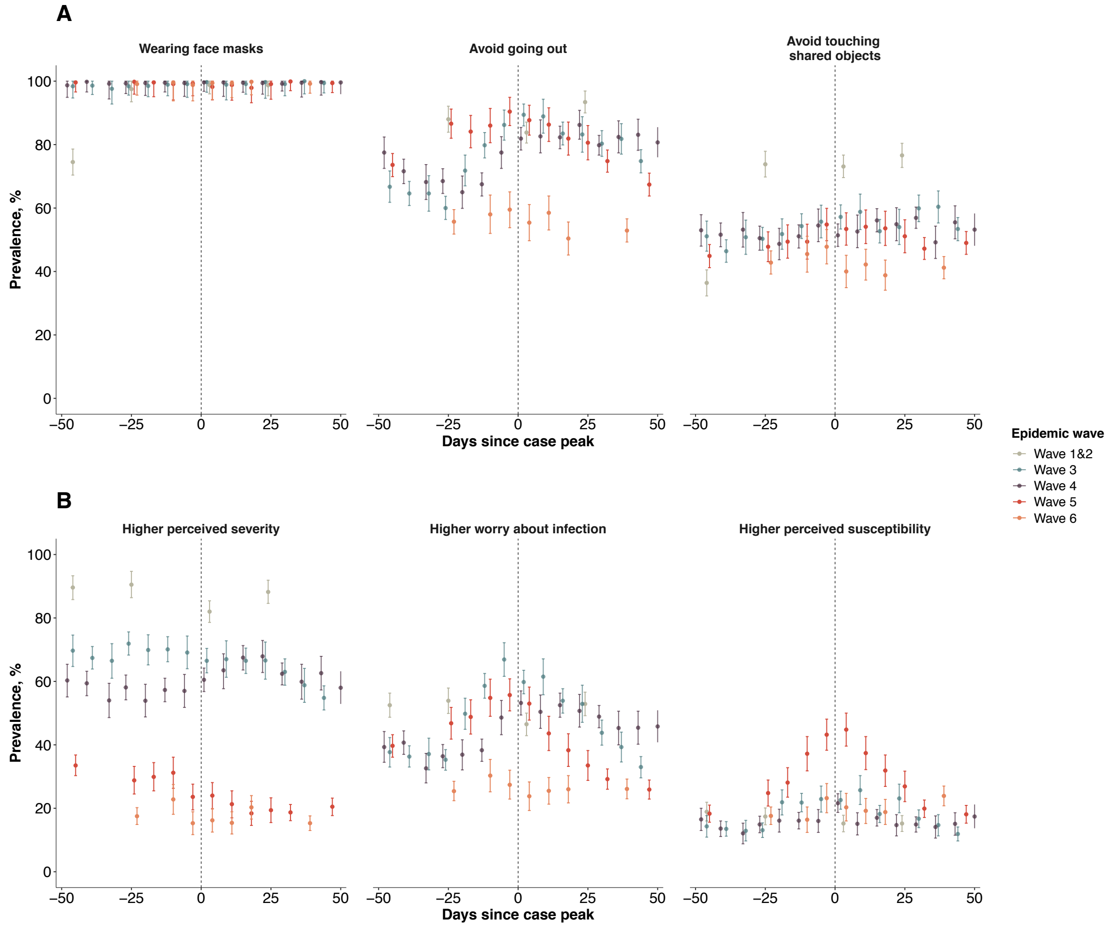

**Figure S8.** Changes in population behaviours related relative to pre-pandemic levels, stratified by waves in Hong Kong. The case peak was determined as the day with the largest number of cases in each wave. Point estimates (points) and 95% confidence intervals (vertical segments) were estimated for each survey.

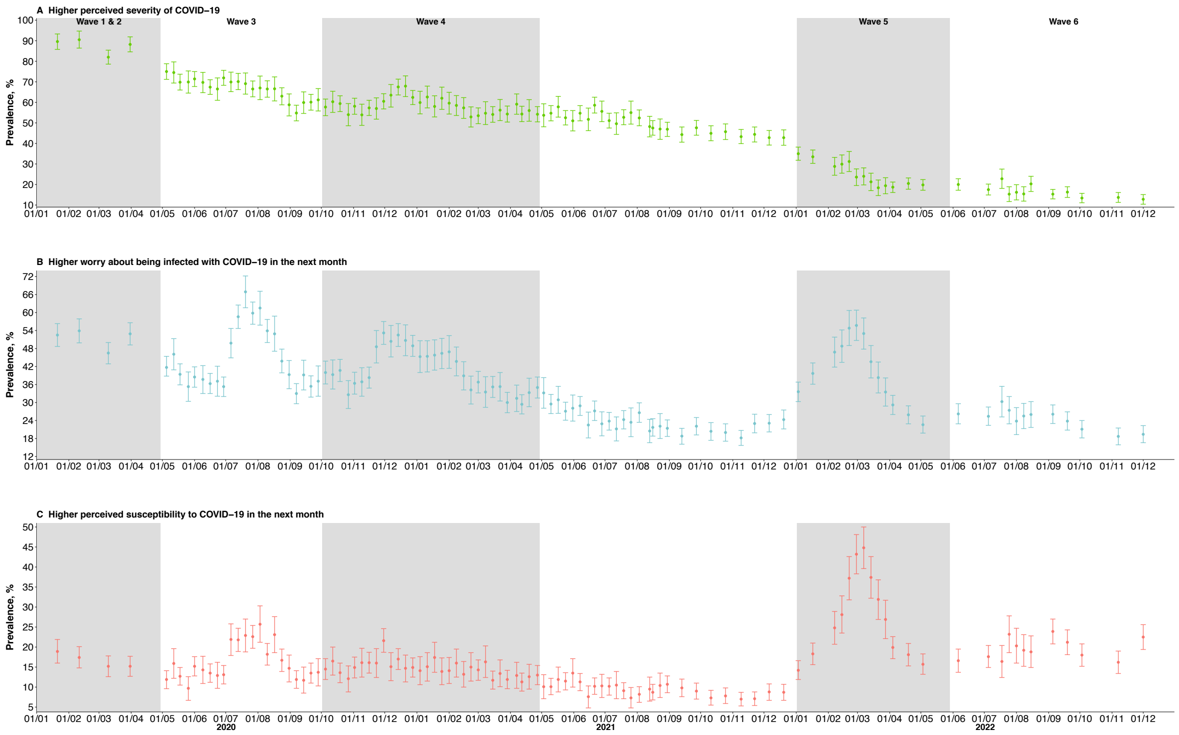

**Figure S9.** Changes in population perceived risks of COVID-19 in the next month among adult population in Hong Kong. Point estimates (points) and 95% confidence intervals (vertical segments) were estimated for each survey. (**A**) Higher perceived severity of COVID−19. (**B**) Higher worry about being infected with COVID−19 in the next month. (**C**) Higher perceived susceptibility to COVID−19.

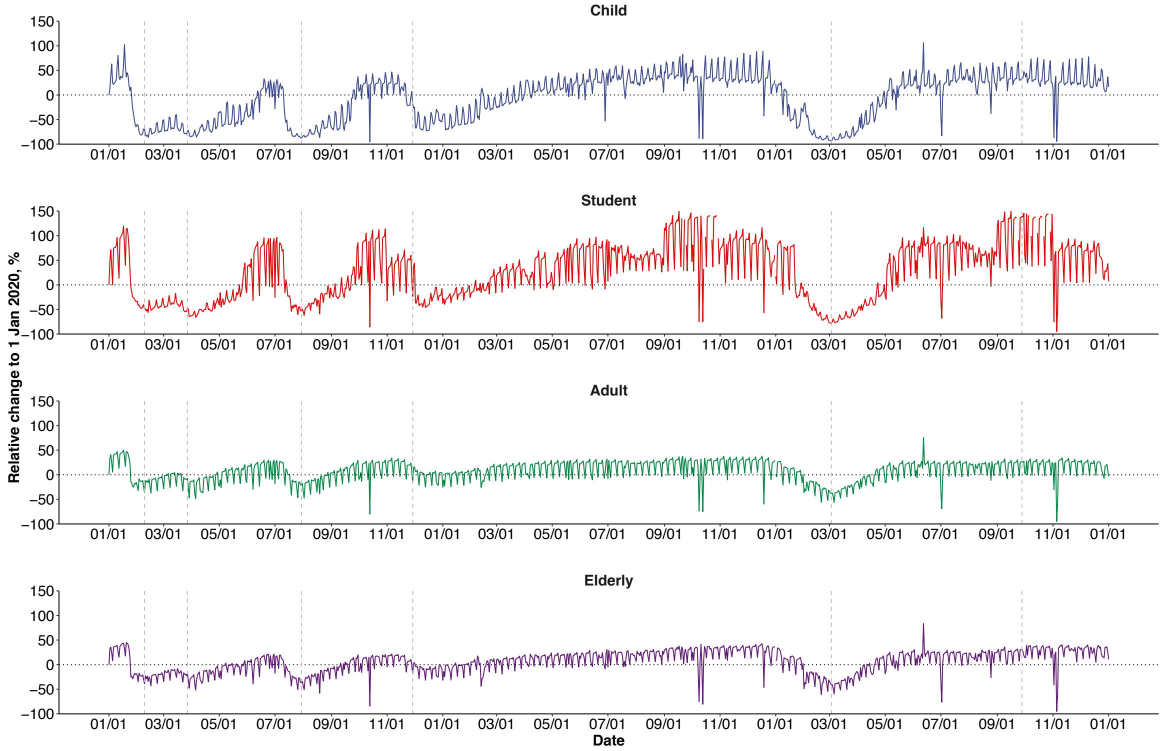

**Figure S10.** Percentage changes in transport transactions using Octopus cards relative to 1 January 2020, stratified by age groups. Vertical dashed lines indicate case peaks in wave 1 to 6.

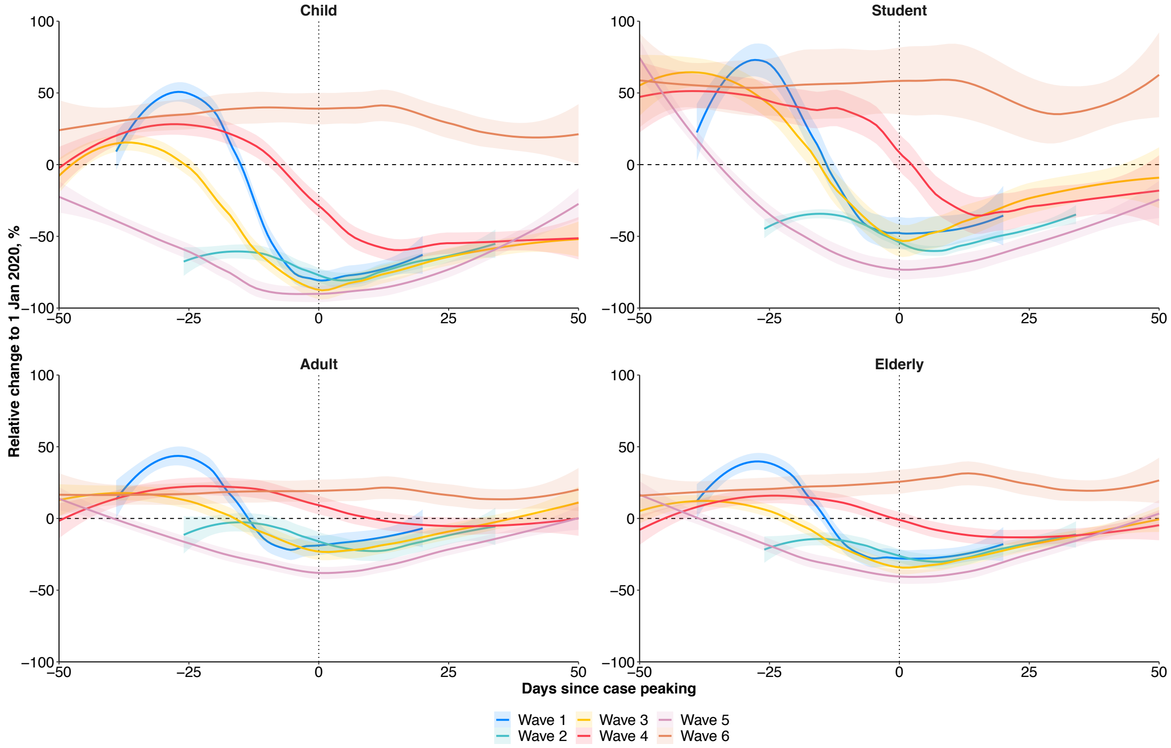

**Figure S11.** Percentage changes in transport transactions using Octopus cards relative to case peak of each wave, stratified by age groups.

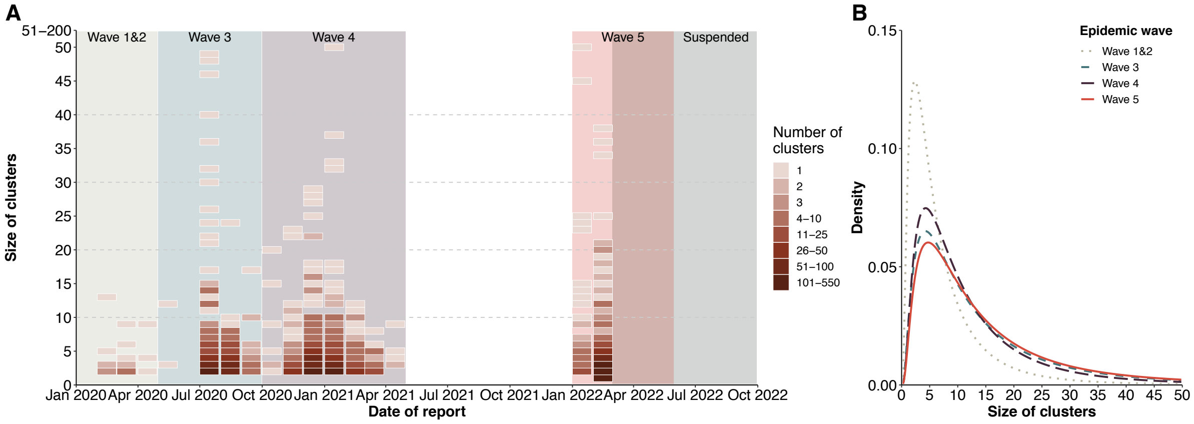

**Figure S12.** Case clustering across different waves in Hong Kong. (**A**) Monthly distribution of the size of clusters that were initiated by index local cases. The grey shade indicates the period when contact tracing, and, thus identification of case clustering were suspended. (**B**) Density distribution of the size of clusters identified across each wave. Log-normal distribution were fitted.

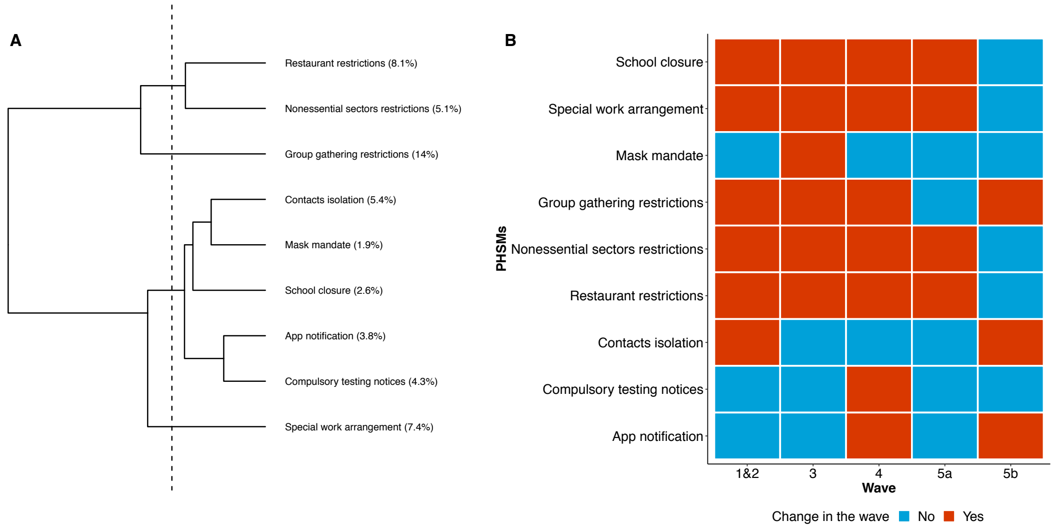

**Figure S13.** Clustering in time of non-pharmaceutical interventions (NPIs) (**A**) and adjustment in NPIs during each wave (**B**) in Hong Kong.

#### 1 Categorization of COVID-19 cases in this study

All probable and confirmed COVID-19 cases identified in Hong Kong were originally reported by the Department of Health as imported cases, close contacts of imported cases, unlinked local cases, close contacts of local cases, possibly local cases or close contacts of possibly local cases.

An imported case refers to a COVID-19 case in a person who has been infected outside of Hong Kong, while a close contact of an imported case is a confirmed COVID-19 case infected locally but identified as a close contact of an imported case. An unlinked local case is a case with no travel history outside Hong Kong within 14 days before the onset of symptoms or confirmation of infection, and whose infection cannot be attributed to any other confirmed case(s). A close contact of local case is a case identified in a cluster and linked to a case infected in Hong Kong. A small proportion of COVID-19 cases in Hong Kong were classified as “possibly local cases”. These were patients who had a travel history to places outside Hong Kong during the incubation period, but the epidemiologic investigation could not determine whether the infection occurred locally after arrival in Hong Kong or whether it had occurred before arrival. Close contacts of possibly local cases refer to confirmed cases linked to a “possibly local” case.

In the current study, we reclassified all the COVID-19 cases confirmed in Hong Kong into six categories: sporadic imported cases, imported index cases, linked-imported cases, sporadic local cases, local index cases, and linked-local cases. We did this by considering their potential contributions to local transmission of COVID-19 and the possible impact on them of different interventions implemented in Hong Kong.

Clusters were defined in this study if one or more cases were linked to another confirmed case ($N \geq 2$). An index case refers to the case first identified in a cluster. For details on case classifications, see Table S2.

From 7 February 2022 onwards (i.e., wave 5b-c), contact tracing and case classifications were largely suspended. From 26 February 2022 onwards, local individuals who tested positive using rapid antigen tests (RAT) were also recorded as confirmed cases via an online self-report system. Since March 2022, inbound travellers who tested positive using RAT during mandatory quarantine were also recorded as confirmed cases.

### 2 Case finding and contact tracing

#### 2.1 Case isolation

From the start of the epidemic until early February 2022, all suspected and confirmed COVID-19 cases were isolated in designated public hospitals for 14 days (or 21 days if infected with emerging variants), regardless of whether symptoms were presented after infections. From February 2022 onwards (i.e., wave 5b and 6), COVID-19 cases confirmed by RT-PCR or RAT were isolated at designated hospitals, community isolation facilities, or homes, depending on their risks of developing severe disease and household environments. Confirmed cases underwent 14 days isolation, which could be shortened to 7 days if the individual had received at least two doses of vaccines and had two consecutive negative results of RAT on day 6 and 7.

#### 2.2 Case finding and laboratory testing

##### 2.2.1 Inbound travelers

Given increases in the numbers of cases among arriving travellers particularly in March 2020, from 25 March onwards, non-Hong Kong residents visiting overseas countries have been barred from entry, and all flight transit services were suspended in Hong Kong while Hong Kong residents were required to undergo 14-day compulsory home quarantine when arriving Hong Kong. From 8 April 2020, all (asymptomatic) inbound travellers by plane were requested to provide deep throat saliva samples for test of SARS-CoV-2 upon arrival.

##### 2.2.2 Screening tests for COVID-19

Laboratory testing initially focused on cases with relevant clinical presentations as well as travel history to COVID-19 affected areas, and was expanded from the end of January 2020 to all inpatients with pneumonia regardless of travel history (i.e., inpatients with pneumonia), and then expanded to all outpatients with relevant clinical presentations in February 2020 (i.e., outpatients). From September 2020 and onwards, screening tests of COVID-19 prior to admission to hospitals were required for all inpatients, regardless of the presence of respiratory symptoms.

##### 2.2.3 Active case finding from high risk groups

Individuals who were considered as having a higher risk of exposure according to their occupations (e.g., staff at Hong Kong International Airport, residential care homes for the elderly and the disabled, nursing homes, Chinese medicine clinics, district health centres, in frontline of franchised bus companies, supermarkets and post, container terminals and fishermen) or places that they have visited, were issued with regular compulsory testing notices. In addition, building-wide compulsory testing notices were issued when there was any confirmed COVID-19 case in the residential building, and all residents were required to undergo RT-PCR tests, with details are described elsewhere.[4]

Self-paid voluntary RT-PCR tests for COVID-19 for symptomatic individuals were also available at community test centres and health care institutions throughout the pandemic. Testing was free for individuals with symptoms, or those without symptoms but required to test under a compulsory testing notice.

##### 2.2.4 Special testing programs

On 1 Sep 2020, the Hong Kong government initiated a 2-week Universal Community Testing Programme (UCTP), aiming to identify hidden cases in the community. In total, 42 cases were identified through the UCTP by testing 1.7 million specimens, with details described in previous study.[5]

In May 2021, the Hong Kong government initiated two rounds of compulsory RT-PCR tests for COVID-19 for all foreign domestic helpers, in response to the identification and suspected transmission of N501Y variant among this segment of the population. In total 340,000 individuals were tested for two rounds (corresponding to very high compliance), among which eight were test positives.

Rapid antigen tests were not widely used in the first two years of the pandemic. However in response to the large fifth wave of Omicron, the Hong Kong government distributed over 17 million rapid antigen tests kits and called for a 3-day daily rapid antigen tests test at home during 8 to 10 April 2022 (https://www.info.gov.hk/gia/general/202204/06/P2022040600547.htm).

#### 2.3 Contact tracing and quarantine

From January 2020 through to early February 2022 (i.e., wave 1 to 5a), diagnosis of a COVID-19 case triggered a contact-tracing investigation to identify other connected cases and possible chains of transmission. A close contact of a COVID-19 case is a person who lived with a case or had a face-to-face interaction for more than 15 minutes when both of them were not wearing face masks. Non-close contacts who had brief contact with the case but not satisfying the definition of close contacts are defined as other contacts. The earliest confirmed COVID-19 case (index case) who triggered the contact tracing investigation might not be the one initiating the transmission in the cluster of cases if earlier infected cases were subsequently detected, i.e. the index case might not be the primary case in a cluster. Contact tracing efforts for non-household contacts were largely suspended from 7 February 2022 onwards.

From January 2020 through early February 2022, the government has repurposed several facilities including holiday camps and hotels to quarantine asymptomatic people who are either close contacts of confirmed COVID-19 cases or travellers from areas with a high-risk transmission of COVID-19, or who violated the order of mandatory home quarantine issued by the Department of Health, or who might not be able to arrange home quarantine after returning Hong Kong from overseas. All asymptomatic close contacts of COVID-19 cases underwent medical surveillance at the quarantine centres for 14 days (or 21 days for people may have been exposed to emerging variants), and a RT-PCR test during quarantine and/or before completion of quarantine, or sent to hospital for testing and management when acute respiratory symptoms were reported. Other contacts will be put under medical surveillance for 14 days and they were sent to hospital for testing and management when acute respiratory symptoms were reported.

During February to December 2022 (i.e., wave 5b and 6), household contacts of COVID-19 cases confirmed by RT-PCR or RAT were required to quarantine at designated quarantine facilities or homes, depending on the transmission risks and severe risks of other household members. Close contacts underwent 14 days quarantine, which can be shortened to 7 days for individuals who had received at least two doses of vaccines and had two consecutive negative results of RAT on day 6 and 7.

#### 3 Population responses to community measures

#### 3.1 Cross-sectional telephone surveys

We conducted a series of cross-sectional telephone survey from January 2020 until the time this study was performed (May 2023), with detailed methods described in previous studies. [6] In each survey, we randomly sampled adults residing in Hong Kong using random-digit dialling at both working and non-working hours. The land-based telephone and mobile phone numbers were generated at a ratio of 1:1. The sample size of each survey was around 500 when conducted weekly and around 1000 when conducted bi-weekly, with detailed calculation methods reported elsewhere. [6] We used rim-weighting to adjust the participants' age, sex, education level, and occupation status distributions to the adult population in Hong Kong, to ensure the sample representativeness.

The participants provided consent and were asked questions about their face mask usage, hygiene and social distancing behaviours in the past 7 days before the survey. For "face mask usage when going out" and "avoid touch or use protective measures with shared objects", we classified the participants as compliant if they responded with "always" or "often" (i.e., half of the time or more); otherwise, they were defined as not compliant (i.e., responded with "never", "sometimes"). We defined the participants as avoiding going out as much as possible if they responded the question with "yes" over "no".

#### 3.2 Octopus usage data reflecting population mobility

We obtained local mobility data from Octopus stored value cards, which are ubiquitous for transport and retail transactions (https://www.octopus.com.hk/en/consumer/index.html). According to the company, 98% of Hong Kong's population aged between 16 and 64 years old possess Octopus cards. We measured the mobility using changes in daily transport transaction relative to the pre-pandemic period (i.e., 1 January 2020).

There are tour types of cards, which mostly depend on the age of the cardholders:

1) Child, age 3 to 11 years old
2) Student, for all primary and secondary school students and university students aged 25 years and below
3) Adult, for adults aged 18 and 64 and who are not students
4) Elderly, for those aged 65 years and above

When calculating overall mobility, we weighted mobility by the age-specific population in Hong Kong.

### 4 Estimation of the time-varying effective reproductive number ($R_t$)

#### 4.1 Model for estimation of $R_t$

Since travel-based measures would directly impact on infections identified from inbound travellers through medical observation or quarantine while community-based measures were assumed to affect occurrence of local infections, the infectiousness of imported cases and local cases were unlikely to be equal. Therefore, we extended the approach described by Cori et al. [7] to estimate the $R_t$ for imported and local cases.

In this approach, we modelled the transmission by a Poisson process. Denote $w_s$ as the probability distribution describing the average infectiousness profile after infection. Then, the rate for infection at time step *t-s* generates new infections in time step *t* is equal to $R_t w_s$, where $R_t$ is the time-varying effective reproductive number at *t*. We use the same idea to separately estimate $R_t$ for imported cases and local cases as follows:

Denote $Y_j(k)$ the number of infection at day *k* for type *j* (1: imported, 2: local case linked with imported cases, 3: local case linked with other local cases). Then, we have:

$$Y_2(t) \sim Poisson\{R_I(t) \sum_{k=1}^{t-1} Y_1(k) w_I(t-k)\}$$

$$Y_3(t) \sim Poisson\{R_L(t) \sum_{k=1}^{t-1} \{Y_2(k) + Y_3(k)\} w_L(t-k)\}$$

where $R_I(t)$ and $R_L(t)$ the time-varying effective reproductive number of imported cases and local cases at time *t* respectively.

We used the smoothing method as in Cori et al., meaning that the transmissibility is assumed constant over a time period $[t-\tau+1, t]$, where $\tau$ is the smoothing parameter. Hence likelihood at a time period *t* is

$$P\left(Y_2(t), \ldots, Y_2(t-\tau+1), Y_3(t), \ldots, Y_3(t-\tau+1) \middle| Y_2(1), \ldots Y_2(t-\tau), Y_3(1), \ldots Y_3(t-\tau)\right)$$

$$= \prod_{s=t-\tau+1}^{t} \frac{\left(R_L^\tau(t)\Phi_L(s)\right)^{Y_3(s)} e^{-R_L^\tau(t)\Phi_L(s)}}{Y_3(s)!} * \frac{\left(R_I^\tau(t)\Phi_I(s)\right)^{Y_2(s)} e^{-R_I^\tau(t)\Phi_I(s)}}{Y_2(s)!}$$

where $\Phi_L(t) = \sum_{k=1}^{t-1}\{Y_2(k) + Y_3(k)\} w_L(t-k)$ and $\Phi_I(t) = \sum_{k=1}^{t-1} Y_1(k) w_I(t-k)$. The total likelihood is the product of time period in the observed data, excluding for the first $\tau - 1$ days due to $\tau$-day smoothing.

##### 4.2 Inference of epidemic curve by infection date

Estimating $R_t$ from the epidemic curve should be based on infection times but we only observed the epidemic curve by symptom onset times.[8] Therefore, we used the deconvolution approach described by Becker et al. [9] to obtain the epidemic curve by infection time from the epidemic curve by onset time, with a given distribution of delay from infection to report. We summarize the approach as the following.

Given a known distribution of the delay from infection to report $f_{delay}(.)$, where

$$f_{delay}(u) = P(delay\ from\ infection\ to\ report = u)$$

Denote $Z_j(k)$ the number of onset at day $k$ for type $j$ (1: imported, 2: local case linked with imported cases, 3: local case linked with other local cases). Then, we have:

$$E\left(Z_j(k)\middle|Y_j(1),\ldots,Y_j(k)\right)=\sum_{u=1}^{k-1}Y_j(u)*f_{delay}(k-u)$$

Denote $\mu_k = E(Z_j(k))$ and $\lambda_k = E(Y_j(k))$, we have

$$\mu_k=\sum_{u=1}^{k-1}\lambda_u*f_{delay}(k-u)$$

Under the assumption that $Y_j(k)$ are independent Poisson variates, $Z_j(k)$ are also independent Poisson variates. Hence, the likelihood function is

$$\prod_k\left[\sum_{u=1}^{k-1}\lambda_u*f_{delay}(k-u)\right]^{Z_j(k)}\exp\{-\sum_{u=1}^{k-1}\lambda_u*f_{delay}(k-u)\}$$

#### 4.3 Assumption on input parameter in data analysis

For incubation period distribution, we use the estimate mean 5.2 days (SD 3.9) estimated from Li *et al.*[10]. Based on the empirical distribution of delay from onset to report, we can construct the distribution for the delay from infection to report by convolution of incubation period distribution and the empirical distribution of delay from onset to report.

The infectiousness since infection $w_t$ was a convolution of incubation period and the infectiousness relative to onset (allowed to be pre-symptomatic) based on viral shedding data, with a shifted gamma distribution [11].

To account for the fact that imported cases did not contribute their entire infectiousness in Hong Kong, we assumed they started to contribute their infectiousness at onset in

Hong Kong (the median of the difference between arrival date and onset date is 0 based on the "eCOVID-19" data). Hence, the infectiousness profile of local cases, $w_L(t) = w(t)$. For the infectiousness profile of imported cases, we assumed that it started at 5 days after infection since the mean incubation period is 5 days. Therefore, the infectiousness profile of imported cases is:

$$w_I(t) = I\left(t \geq 5\right) * \frac{w(t)}{\sum I(t \geq 5)w(t)}$$

We analyse the epidemic curve up to Sep 30, 2020, and take $\tau = 14$ in our analysis, to avoid too bumpy estimates for time-varying reproductive number.

In the dataset, local cases were further classified as unlinked local cases or local cases linked with other local cases. In our analysis, we assumed all unlinked local cases were infected from other local cases due to strong control of imported cases like 14-day quarantine on all inbound travellers.

Due to the fact that not every infection would be identified particularly in the beginning of an emergency infectious disease outbreak[12], we assumed that there were 15 undetected cases in the beginning of the third wave (Jun 11 – 23, 2020). Without such modification, the $R_t$ estimates could be >10 in the beginning of the third wave, which is unrealistic.

##### 4.4 Inference

To inference $R_t$, we first use the deconvolution summarized in Section w.2. An EM algorithm with a smoothing step is also included to obtain a smoother epidemic curve to enhance interpretation. This is achieved by using the backprojNP function in the surveillance package in R.

After obtaining the epidemic curve by infection time, we use the model in Section 4.1 to estimate $R_t$ for local cases and imported cases. We use a Markov chain Monte Carlo approach with standard Metropolis-Hastings algorithm to estimate the model parameter. The is achieved by using the Rcpp package with self-written cpp function, which is available at https://github.com/timktsang.

We accounted for the uncertainty of input parameter, including incubation period and infectiousness profile to obtain the final estimates of $R_t$ in addition to model parameter uncertainty by following the approach described by Salje et al.[13] We reconstructed 1000 epidemic curves. For each epidemic curve, we first sample the parameters for the incubation period distribution. Then we used the deconvolution with that incubation period distribution and estimate $R_t$ for local cases and imported cases. Then we resampled the date of infection for the 15 undetected cases in the beginning of the third wave. Then we presented the mean, 2.5% and 97.5% quantiles for those 1000 $R_t$ estimates for each time point.

#### 5 Estimation of potential effect of NPIs

To determine the individual NPIs included in the package, we assessed the intervention clustering patterns, the variance of transmissions explained by each intervention, and policy adjustments during each wave.

First, we fitted a hierarchical clustering model to all community-based NPIs, allowing for home quarantine for inbound travellers (as this aimed to reduce mixing with the population in the community) (Figure S1). We also fitted univariable log-linear regression to daily $R_t$ and each of the above NPIs, to examine the variance of transmission explained by each NPI. We also looked at whether there were any changes in the stringency of each individual NPIs during each wave.

Based on the results from the abovementioned analyses (Figure S1), we included six NPIs to parsimoniously reflect the measure changes over time, namely restrictions on restaurant operations, restrictions on group gathering, school closure, compulsory testing notices,  quarantine at home and work-from-home arrangement. The impact of NPIs wa estimated using the following equation:

$$log(R_{t,w}) = log\left(R_{0,w}\right) + \sum_{w \in W} \sum_{t \in T} \alpha_w Z_{w,t} + \sum_{j \in J} \sum_{t \in T} \beta_j X_{j,t} + \epsilon_t$$

where:

- $R_{t,w}$ denotes the reproduction number at time $t$ ($t \in T$) in wave $w$ ($w \in W, where\ W = \{1, 2, 3, 4, 5a\}$)

- $R_{0,w}$ denotes the initial transmissibility in the beginning of each wave, which may be affected by the basic reproduction number of the dominant strains and the population susceptibility
- $\alpha_w$ denotes the wave-specific coefficient for work-from-home arrangement $Z_{w,t}$
- $\beta_j$ denotes the coefficient for each of the six included NPIs $X_{j,t}$ at time $t$

With the estimated coefficients of NPIs, we derived the expected Rt for each wave (i.e., $R_{0,w}$) under different PSHMs packages (Figure 2A).

All analyses were conducted in R version 4.0.0 (R Foundation for Statistical Computing, Vienna, Austria).

#### 6 COVID-19 vaccines

Two COVID-19 vaccines were available in Hong Kong, the inactivated vaccine CoronaVac (Sinovac) vaccine and the mRNA vaccine BNT162b2 (BioNTech/Fosun Pharma/Pfizer). These were offered for free starting in late February 2021, with third and fourth doses available for adults starting from November 2021 and early April 2022, respectively. Individuals were able to choose which vaccine to receive, but the type of vaccine used for the first two doses should be the same based on manufacturer's drug insert and expert recommendations. Starting from the third dose, heterologous doses were permitted.

We obtained data on population vaccine uptake, where vaccine doses and types were tracked daily and stratified by individual age group. We obtained data on population by age from the 2021 Hong Kong population census. We calculated the overall proportion of the population fully vaccinated as the number of individuals who received at least two doses of COVID-19 vaccines divided by the corresponding population in Hong Kong, stratifying by wave and trajectory. We also calculated the proportion fully vaccinated among the elderly as the number of individuals aged ≥65 years who received at least two doses of COVID-19 vaccines divided by the total population of people aged ≥65 years in Hong Kong.

We obtained vaccine history in among COVID-19 cases from the Department of Health. Although cases may receive vaccines other than Sinovac and BNT162b2 outside Hong

Kong, we found that among COVID-19 cases who received at least one dose, 98% of them received Sinovac and/or BNT162b2.

#### 7 Oral antiviral drugs against COVID-19

The Hong Kong Hospital Authority began prescribing molnupiravir (Merck) from 26 February 2022 and nirmatrelvir/ritonavir (Pfizer) from 16 March 2022 to COVID-19 patients who were considered to be at higher risk of severe disease, following a standard treatment protocol.[14] Both antiviral drugs were equally available across public hospitals. We obtained data on antiviral drug prescriptions from the Hospital Authority, including the first date when each case receiving molnupiravir and nirmatrelvir/ritonavir, respectively. We linked data on antiviral drug usage to the COVID-19 case database obtained from the Department of Health, to obtain data on demographics and vaccine histories of these cases.

We first examined the antiviral usage among the elderly by calculating the proportion of COVID-19 cases who were aged ≥65 years and received molnupiravir, nirmatrelvir/ritonavir or both among all COVID-19 cases aged ≥65 years during the same wave and trajectory. We also repeated the analysis restricting to COVID-19 cases aged ≥65 years who did not receive COVID-19 vaccines (i.e., unvaccinated adults aged ≥65 years).

#### 8 Mortality rate and case-fatality risks of COVID-19

To characterize the temporal change of disease burden associated with COVID-19, we estimated the overall mortality rate as the number of deaths of COVID-19 cases divided by the population in Hong Kong and the time of across wave and trajectory (Table S1). We repeated the analysis to calculate the mortality rate for the elderly (i.e., aged ≥65 years), where we obtained the corresponding population denominator of adults aged ≥65 years from the Hong Kong Population census.

We calculated the mortality rate among the unvaccinated population as the number of fatal COVID-19 cases who did not receive COVID-19 vaccines divided by the unvaccinated population-time, which was estimated as the product of population in

Hong Kong, the proportion of population receiving no vaccines and time observed in each wave and across trajectories.

Similarly, we calculated the mortality rate among vaccinated adults aged ≥65 years as the number of fatal COVID-19 cases among individuals aged ≥65 years who did not receive COVID-19 vaccines, divided by the population-time of fully vaccinated adults aged ≥65y. The fully vaccinated adults aged ≥65y was estimated by taking the product of the population aged ≥65 years in Hong Kong, the coverage of at least two doses among aged ≥65 years, and the relevant duration of each wave and trajectory period.

We calculated case-fatality risks as the number of COVID-19 deaths divided by the total number of all confirmed COVID-19 cases during each wave and trajectory period. We also conducted separate analyses focusing on different groups: all adults aged ≥65 years, vaccinated adults aged ≥65 years with at least two doses, and unvaccinated individuals of all ages.
